# Haploinsufficiency, *de novo* and biallelic missense variants in *CSMD2* are associated with neurodevelopmental disorders

**DOI:** 10.64898/2026.09.24.26361611

**Authors:** Clara Pailler-Pradeau, Marianne Victoria Lemée, Caterina Cevallos, Francesca Mattioli, Jacqueline Chrast, Sissy Bassani, Hossein Darvish, Ginevra Zanni, Giovanna Stefania Colafati, Lorena Travaglini, Francesco Nicita, Shivarajan M. Amudhavalli, Caitlin Lawson, Joseph G. Gleeson, Isabella Stuewe, Liana Friedman, Maha S. Zaki, Lauren Bell, Elise Boucher Brischoux, Paul Kuentz, Nico Fuhrmann, Victoria Lampkemeyer, Christian Netzer, Thomas Smol, Catherine Vincent-Delorme, Zafar Iqbal, Mathias Toft, Tarab Zehra, Sidra Kaleem Jafri, Ambrin Fatima, Mukhtar Ullah, Zainab Akhtar, Tayyaba Shan, Sabika Firasat, Saghar Ghasemi Firouzabadi, Abbas Tafakhori, Sarina Sharbatkhori, Faezeh Jamali, Maximilian Balbach, Friedhelm Hildebrandt, Emily O’Heir, Lynn Pais, Alba Sanchis-Juan, Michael E. Talkowski, Quinten Waisfisz, Lotte Kleinendorst, Giovanna Ambrosini, Trisha Kuchta, Zoltan Kutalik, Nicolas Guex, Stylianos E. Antonarakis, Muhammad Ansar, Christelle Golzio, Alexandre Reymond

## Abstract

CUB and Sushi domains-containing CSMD proteins are involved in neuron development. Variants in the *CSMD1-3* paralogs were GWAS-associated with cognition, attention-deficit hyperactivity disorder (ADHD), schizophrenia and brain structure, suggesting that these genes could be linked to neurodevelopmental disorders (NDD). Indeed, rare biallelic variants in all three *CSMD* paralogs were recently associated with epilepsy, while rare biallelic variants in *CSMD1* were also linked with intellectual disability and cortical malformations. We identified nine individuals from eight families carrying *CSMD2* biallelic missense variants in developmental delay/intellectual disability cohorts. Affected individuals presented with autism spectrum disorder (ASD), ADHD and brain anomalies. Consistent with gnomAD metrics that suggest *CSMD2* haploinsufficiency, we also report two families segregating monoallelic *CSMD2* frameshift variants, which complement a described *de novo* truncation variant in an ASD proband. We concomitantly report two phenotypically overlapping individuals with *de novo CSMD2* missense variants and compare them to twelve *de novo CSMD2* cases identified in published trio analyses. UK Biobank analyses showed nominally significant enrichment of ADHD, ASD and neuroticism in volunteers carrying at least one rare *CSMD2* variant. *In silico* modelling and immunofluorescence assays showed that mutated residues in affected individuals with biallelic and *de novo* missense variants are clustering in two regions and that the encoded proteins showed impaired membrane localization and an increased cytoplasmic aggregation.

Zebrafish crispant larvae for the orthologous *csmd2* presented with microcephaly, smaller cerebellum, and reduced Purkinje cell size. Larvae also displayed decreased swimming velocity, diminished connectivity between the optic tecta, and abnormal peripheral neuronal branching.

We demonstrate that rare, likely pathogenic variants in *CSMD2* are associated with both autosomal recessive and dominant forms of NDD further substantiating the functional significance of CSMD paralogs in neurogenesis/synapse formation.

## INTRODUCTION

CUB and Sushi Multiple Domains proteins (CSMD) are encoded by three highly similar paralogs in mammals, *CSMD1*, *CSMD2* and *CSMD3* (60-65% identity at the encoded amino acid level) as well as a strong evolutionary conservation across vertebrates^1,2^ (**Figure S1, Figure S2**) mainly expressed in the brain^3^. They are made of an alternation of fourteen CUB and fourteen Sushi domains, followed by a maximum of fourteen contiguous Sushi domains, depending on the isoforms, a transmembrane domain and a short intracellular C-terminal tail^4^. The 110 residue-long CUB domains are often found in developmentally-regulated extracellular or membrane-associated proteins, such as neuropilins, involved in the formation of neuronal circuits^5^, while the 60 amino acid long Sushi domain, also known as short consensus repeat (SCR) or complement control protein (CCP) module, contains four invariant cysteines that form two disulfide bridges. Sushi domains are involved in recognition and binding processes and are, for example, found in proteins of the complement cascade and in the coagulation factor XIII B chain^6^. The complement cascade role in synaptic plasticity and pruning through tagging of obsolete synapses was shown to be important for cognitive function^7^.

*CSMD2* is expressed in the human brain and the cerebellum with a peak of expression level just before birth, a pattern of expression conserved through various vertebrate species^8^. Mouse *Csmd2* is expressed in excitatory projection neurons and inhibitory interneurons with a somatodendritic pattern^9^. The encoded protein is localized at synapses of the neocortex, the hippocampus and the retina and was shown to interact with synaptic scaffold protein PSD95, an interaction necessary for its synaptic localization^9^. PSD95 is encoded by *DLG4*, which is linked to the autosomal dominant intellectual developmental disorder 62 (MRD62; MIM#618793)^10,11^. *Csmd2* is important for dendrite and dendritic spine development as its inactivation resulted in reduced dendritic complexity and a decrease in the number of dendritic spines^9^. Mouse *Csmd3* was similarly shown to play a role in dendritic development and maintenance^12^, while assays with human stem cell-derived neurons lacking *CSMD1* suggested that it regulates synapse elimination during development^13^. Consistent with a role in neuronal development, common variants mapping to the *CSMD1* and *CSMD2* loci have been associated by GWAS (genome wide association studies) with brain/cerebellum structure^14^, cognitive performance, education attainment, depression, schizophrenia^15,16^ and drug/alcohol consumption^17^. The *CSMD2* locus was also associated with attention deficit hyperactivity disorder (ADHD)^18,19^, whereas *CSMD1* and *CSMD3* were linked to cyclothymia/bipolar disorder, post-traumatic stress disorder (PTSD) and chronotype/insomnia and modification of the reward response, Tourette syndrome and depression, respectively^12,20–30^. *CSMD1* and *CSMD2* were associated to schizophrenia through gene expression changes of immunity-related processes involved in complement activity and antigen presentation that influence long-term potentiation of synaptic strength, which is important for learning and memory^16^. Further substantiating the importance of the CSMDs in neuronal development, bi-allelic variants in the three *CSMD* paralogs were recently associated with epilepsy^31,32^ and in *CSMD1* with intellectual disability (ID) and cortical malformations^20,33^.

We describe thirty-two individuals presenting with NDD and brain anomalies carrying rare, likely pathogenic variants in *CSMD2*. *In silico* 3D modelling and protein localization assays suggest functional impairment of the identified variants. Crispant zebrafish larvae ablated for the *CSMD2* ortholog showed both central and peripheral nervous system perturbations.

## MATERIALS AND METHODS

### Samples

Signed informed consent forms were obtained from all affected individuals participating in this study or their guardians. The current study was approved by the respective IRBs, e.g. the CER (Commission d’éthique de la recherche) of the canton of Vaud (Protocol number: CER-VD 2021-01400). Whole exome sequencing was performed as previously described^34^.

### 3D model

The *in silico* 3D contextualization of CSMD2 mutations as well as their predicted effect were established using the Swiss-Pdb Viewer^35^, the UniProt entry Q7Z408 annotation and its corresponding alpha-fold model. Since for long proteins, alpha-fold models are only provided as overlapping section of 1400 residues, we applied the following procedure to obtain a full-length model: alpha-fold fragments AF-Q7Z408-F1-model_v2.pdb, AF-Q7Z408-F7-model_v2.pdb and AF-Q7Z408-F12-model_v2.pdb were loaded in Swiss-PdbViewer and residues renumbered to match the Uniprot entry using the “Rename current layer” option. Then the section AF-Q7Z408-F7-model_v2.pdb was superposed onto the section AF-Q7Z408-F1-model_v2.pdb using the “Fit molecules from selection” option using Carbon alpha atoms of the beta-strand residues Gly1372-Ala1382 (rmsd: 11 atoms @0.2A). Then beta-strand residues Thr2551-Ser2558, Gly2560-Gln2564, Ala2569-Leu2574, Leu2578-Ser2580, and Gln2587-Val2591 of the section AF-Q7Z408-F12-model_v2.pdb were superposed onto AF-Q7Z408-F7-model_v2.pdb (rmsd: 27 atoms @0.18A). Finally, a full-length model was produced by selecting residues [start-Phe1377][Ser1378-Pro2590][Val2591-end] of each respective section with the “Create merged layer from selection” option.

### Immunofluorescence assay for missense variants

Plasmids carrying a N-terminal 3xFLAG-tagged mouse *Csmd2* cDNA, either wild-type (WT) or containing one of the variants identified in the affected individuals (P73S, G85D, P1431L, R1609W, G1632D, Q1765R, R1995Q, K2175M, T2190I, R2445H, A2534T, R2635C, D3141H, S3366C and L3532R) under the CMV promoter were synthesized by VectorBuilder (Chicago, IL, USA). The plasmids were amplified in *E. coli* before being purified using a MidiPreps kit (QIAGEN, Plasmid Plus Midi kit, 12943). HEK293T cells were seeded in 12-well plates at a density of 1.4 x 10^5^ cells/well. Once they reached 60-70% of confluence, they were transfected with plasmids encoding either WT or an affected individual-specific *CSMD2* variant isoform using a Lipofectamine 3000 kit (L3000001, Invitrogen). For each well, the diluted Lipofectamine 3000 reagent was prepared (50 μL of Opti-MEME Medium and 1.5 μL of Lipofectamine 3000 reagent), as well as one master mix of DNA per plasmid (50 μL of Opti-MEM Medium, 1 μg of DNA and 2 μL of P3000 reagent). The diluted DNA mixture was then combined with an equal volume of diluted Lipofectamine 3000 reagent and incubated for 15 min at room temperature. Subsequently, 100 μL of DNA-lipid complex was added to each well, and the cells were incubated overnight. The next day, the cells were fixed using 4% PFA for 15 min at room temperature and permeabilized with Triton 0.25% for 5 min, incubated for 30 min at 37°C with 10% BSA/PBS, before incubation with the primary antibody (monoclonal mouse anti-FLAG M2 F1804 or F3165, Sigma-Aldrich, 1:1000 in 3% BSA/PBS) for 2h at 37°C, and the secondary antibody (anti-mouse, Alexa Fluor 488, Sigma-Aldrich, 1:1000 in 3% BSA/PBS) for 45 min at 37°C in the dark. Coverslips were mounted using the Vectashield, H-1200 mounting solution containing DAPI. Images of the transfected cells were acquired using a Zeiss Axiovision Imager M1 microscope at 100x magnification with a Plan Neofluar objective. We measured the intensity of fluorescence at the membrane, using the Fiji software to calculate the integrated density (IntDen) of fluorescence, which corresponds to the product of the mean gray value and the area^36^. The statistical analysis was performed through Wilcoxon tests on R to compare each condition to the WT (wild type) condition.

### UK Biobank search

To assess whether *CSMD2* associations observed in clinical cohorts extend to the general population, we conducted rare variant analyses in the UK Biobank (UKBB). The UKBB has enrolled about half a million British volunteers from across the United Kingdom^37^ between 2006 and 2010. Its extensive genotypic and phenotypic information, constitutes a powerful resource for elucidating the genetic architecture of health-related traits. We focused on a subset of unrelated individuals of European ancestry (defined as in.white.British.ancestry.subset==1 in the file ukb_sqc_v2.txt) with available whole-exome sequencing data (UKBB field identifier #23149).

Sample-and individual-level, quality control was performed using PLINK v2^38^. Participants were excluded if they exhibited high per-sample missingness (--mind 0.1), mismatching submitted and genetically inferred sex or had withdrawn consent at the time of the analysis. We pruned variants retaining those with minor allele frequency (MAF) < 0.1% (--MAF 0.001) and missingness rate < 10% (--geno 0.1). Predicted loss-of-function (pLoF) variants were identified using Ensembl VEP v95 and restricted to high-confidence variants as indicated by LOFTEE^39^. Missense variants were retrieved from SnpEff^40^ annotations available in the UKBB (ukb23145_300k_OQFE.annotations.txt.gz file) and ranked by deleteriousness using five algorithms in dbNSPF^41^ v3.2 database: SIFT, PolyPhen2 HDIV, PolyPhen2 HVAR, LRT, and MutationTaster^42^. They were classified as high-confidence (5/5), medium-confidence (1/5-4/5), and likely benign (0/5) according to the number of algorithms predicting a deleterious effect. We computed pLoF and missense variant counts using PLINK v2 (--score--no-mean-imputation), and defined carriers as individuals harboring at least one corresponding variant in CSMD2. Binary disease phenotypes were derived from ICD10-diagnoses (UKBB field identifier #41270) and professionally diagnosed mental health problems (UKBB field identifier #20544) encompassing childhood autism (F840), Asperger’s syndrome (F845), epilepsy (G40), intellectual disability (F70, F71, F72, F78, F79), schizophrenia (F20 or code=2), ADHD (code=18), and psychosis (code=3). Additionally, continuous traits, including fluid intelligence (UKBB field identifier #20016) and neuroticism (UKBB field identifier #20127) scores were adjusted for sex, age, age^2^, genotyping batch, and the first 40 principal components. Associations between rare CSMD2 variants and its putative phenotypic outcomes were assessed across four variant categories: pLoF, high-confidence missense, medium-confidence missense, and combined medium and high confidence missense variants, using Fisher’s exact test for the binary traits and a two-sided t-test for the continuous traits. To prevent signal spill-over between variant classes, we excluded missense-only carriers in pLoF analyses, and all pLoF carriers, in missense analyses.

### Zebrafish model

All procedures were performed in accordance with protocols approved by the local ethics Committee (UNIL and IGBMC) and with the approval of the relevant institutional authorities (canton Vaud veterinary office authorization VD-H28; French Ministry of Higher Education and Research authorization APAFIS# #49694-2024050316122698.). Zebrafish (*Danio rerio*, AB strain obtained from the Zebrafish International Resource Center (ZIRC), Oregon) were housed at 28.5°C in Active Blue Racks (Tecniplast, Buguggiate, Italy) with 20 fish per tank maximum, in a light/dark cycle of 14/10h. Adult WT zebrafish were put in a crossing tank the night before, with 2 females and 3 males per crossing tank separated using a divider. On the day of the experiment, the divider was removed, and eggs were collected approximately 10 minutes after laying to have time to be fertilized. Eggs at the one-or two-cell stage were injected with 1nL of a mix containing either 1 μL of single guide RNAs at 100 ng/μL, 1 μL of TrueCut™ Cas9 v2 (Invitrogen) at 200 ng/μL (which were incubated together for 5 minutes before adding the rest), 1 μL of phenol red 6X, 1μL of KCl at 200 mM and 1μL of water (F0 crispant group) or the same mix replacing Cas9 by water (mock group). To engineer F0 crispant larvae, we used a mix of the following four different sgRNA targeting exons 4, 19, 24 and 56 (CTTGTCCCCGATGTTGAAGGTGG; GGATGATTGAGACTTCACATGGG; TCCCGTTTAAAACCGAACTGAGG; TCCGTATGCACAGATCTCAGGGG), designed using the CHOPCHOP tool^43,44^ and ordered from Invitrogen TrueGuide (Synthego, Redwood City, CA, USA). After injection, eggs were incubated for 24h at 28°C in blue water, dead eggs were removed, and half of the water was changed.

The CRISPR-Cas9 efficiency of each sgRNA was determined on 5 dpf larvae through a mismatch detection assay with T7 endonuclease 1 (New England Biolabs, Ipswich, MA, United States). First, DNA was extracted using 15 μL per larva of a mix containing 7 μL of Tris-HCl 1.5 M pH 8.8, 4 μL of EDTA pH8, 2 μL of Triton 100X, 10 μL of protein K at 20 mg/mL and 977 μL of DNase free water. Crispant larvae were incubated in the mix for 2h at 55°C and vortexed regularly, and then at 95°C for 10 min. Then the DNA was amplified using primers (Sigma-Aldrich, Darmstadt, Germany) flanking the sgRNAs target sites (GCCAATAATTTTGACGTGTGTG and CTGTGCATCCAACCTGCTACTA, CCCCTTGGGTAATGATAGGAAT and TGCCGCTGTGTAAAAACTTG, TGCCGCTGTGTAAAAACTTG and CAGTGATGCAGTAGTTGGCTTT, TGTAACCCTGGGTACTACCTGC and AACAGCAATTGTGTGGATGAAC). The mix for the amplification was made of 1 μL of each primer at 10 μM, 3 μL of DNA, 12.5 μL of RedTaq (Sigma-Aldrich, P0982), the program used went from 94°C to 65°C in steps of 30 sec with −0.5°C each time. After that the PCR products were denatured at 95°C for 2 min and through a touchdown of −2°C per second down to 85°C and then −0.1°C per second down to 25°C. This step was followed by the addition of 1 μL T7 endonuclease 1 and an incubation of 15 min at 37°C. After these 15 min, 1.5 μL of 0.25 M EDTA was added to stop the reaction. The samples were then loaded on a 2% agarose gel.

The swimming behavior of 5 dpf crispant larvae was studied using the Zebrabox® recording system (Viewpoint, Lissieu, France) for 15 min in the light and 15 min in the dark with infrared light. Fast velocity was calculated combining two distance categories recorded by the tracking system: small movement distance (smldist, 3–6 mm/s, corresponding to slow/moderate locomotor activity) and large movement distance (lardist, >6 mm/s, corresponding to rapid locomotor activity). Fast velocity = (smldist + lardist) / time; time is the duration (in seconds) of the recording. This metric reflects the total distance covered by movements classified as moderate-to-fast (excluding purely stationary or minimal movement, i.e. below the 3 mm/s threshold), normalized by the recording time to yield a velocity (mm/s). The experiment was repeated four times, and the results were pooled together. The head size was measured between the eyes of the larvae, at the level of the pupils^45,46^. The body length was measured from the mouth to the tail’s tip of the larvae, following the spinal cord. The statistical analyses were performed on R using Wilcoxon tests comparing the mock and the knock-down groups. For parvalbumin 7 (Pvalb7) immunolabelling, larvae were fixed at 4 dpf for 4h in a solution of 4% of PFA in PBS-Triton at room temperature, then washed 10 min in PBS-Triton 0.1%, then bleached for 10-15 min, incubated for 1h at room temperature in a blocking solution (46,5 mL PBS 1X, 500 μL Triton 1%, 500 μL DMSO 1%, 0,5 g BSA 1%, 2,5 mL sheep goat 5%) and then incubated overnight with the primary antibody (mouse anti-Pvalb7 (kindly provided by Dr Masahiko Hibi from the University of Nagoya, Japan, 1:500 in blocking solution) at 4°C. The next day, the larvae were incubated with the secondary antibody (goat anti-mouse Alexa Fluor 488, A11001, Invitrogen, 1:500 in blocking solution) for 2h at room temperature in the dark and stored in PBS-Tween 0.1% at 4°C. For acetylated tubulin (AcTub) immunolabelling, larvae were fixed at 4 dpf overnight at 4°C in Dent’s solution (80% methanol and 20% DMSO). The next day the larvae were rehydrated in 3 solutions with a decreasing amount of methanol and an increasing amount of PBS (75/25, 50/50 and 25/75), before being bleached, treated with proteinase K at 10μg/mL in PBS for 10 min, PFA 4% for 20 min and IF buffer (1% BSA in PBS-Tween 0.1%) for 10 min, incubated in blocking solution (9 mL IF buffer and 1 mL FCS 100%) for 1h, followed by an overnight incubation of the primary antibody (mouse anti-AcTub, T7451 Sigma-Aldrich, 1:1000 in blocking solution). On the 3^rd^ day, larvae were incubated in the dark for 2h with the secondary antibody (goat anti-mouse Alexa Fluor 488, A11001, Invitrogen, 1:500 in blocking solution) and stored in PBS-Tween 0.1% at 4°C. Stained larvae were imaged using a MacroFluo ORCA Flash Leica macroscope and an ORCA Flash 4.0 Hamamatsu Digital C11440 Camera. Pvalb7 immunolabelled cerebella, AcTub immunolabelled heads and tails were acquired at 9.2x, 8x and 5x magnification, respectively before analysis with the Fiji software^36^. The analysis of the motoneurons was performed in the following way, out of 8 motoneurons in the tail of the larvae we counted how many presented an abnormal shape after the immunolabelling directed against acetylated tubulin^47^. The minimum distance between the two optic tecta was measured after the immunolabelling directed against the acetylated tubulin and then normalized to the mean distance of the Mock group. The area of one optic tectum per larva was measured after the immunolabelling directed against the acetylated tubulin and then normalized to the mean tecta area of the Mock group. The number of inter-tecta axonal tracts was counted after the immunolabelling directed against the acetylated tubulin^45,48^. The area of the cerebellum was measured for each larva after the immunolabelling directed against the acetylated tubulin, before being normalized to the mean cerebellum area of the Mock group. The number of Purkinje’s cells in one half of the cerebellum was counted for each larva after the immunolabelling directed against the parvalbumin 7. To determine the mean area of the Purkinje’s cells, we individually measured the area of 10 distinct Purkinje’s cells localized in one half of the cerebellum of each larva, following immunolabelling against the parvalbumin 7, and calculated the mean area per larva. These mean values were then normalized to the mean Purkinje cell area of the Mock group. The head size was measured under a binocular microscope at a 6,3x magnification and the body length at a 2x magnification on the same larvae which were used for the immunolabeling.

## RESULTS

### Inheritance, variants and phenotypes

Within our cohort of exome-sequenced multiplex neurodevelopmental disease (NDD) consanguineous families we identified two affected Iranian brothers (affected individuals’ number 1-2), with homozygous missense variant T2190I in *CSMD2* (MANE NM_001281956.2)(**Figure S3A, Table S1**). Using GeneMatcher^49^ and literature search, we subsequently identified seven more unrelated affected individuals carrying predicted deleterious biallelic variants in that gene either at the homozygous state (affected individuals 3-6) or as compound heterozygotes (affected individuals 7-9, **Table S1**). Affected individual 9 was previously described^31^. Variant predicted deleteriousness and frequencies are detailed in **Table S2**. Consistent with an autosomal recessive disease, no homozygous or compound heterozygous samples with REVEL score ≥0.644 were identified in gnomAD v2.1.1. Of note, the kidney impairment of affected individual 6 could be associated to an additional *NPHP4* variant (MANE NM_015102:c.1149_1150insCC:p.A384Pfs*23) (**Table S1**), which has already been classified as pathogenic in ClinVar (ClinVar Variation ID: 562343). All nine affected individuals present with learning difficulties with eight having been formally diagnosed with ID. Brain MRIs showed brain anomalies in all seven assessed individuals, such as generalized brain atrophy with expansion of the cerebrospinal fluid convexities, rhomboencephalosynapsis, severely dilated cerebral ventricles, retrocerebellar cyst undergoing ventricular cysternostomy or bilateral symmetrical increase in hypodensity of white matter (**Figure 1A** and **Table S1**). The majority of affected individuals presented with

**Figure 1.**
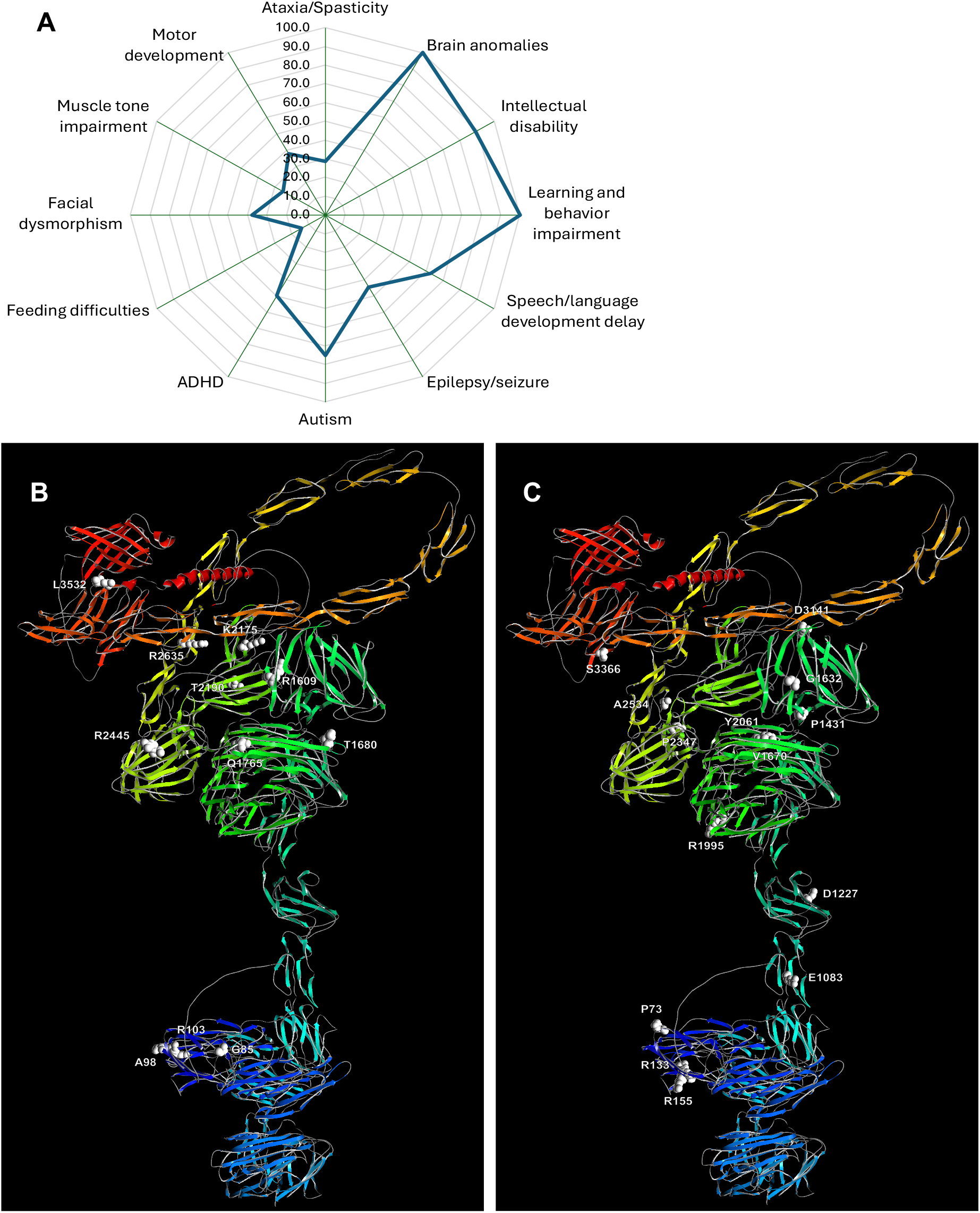
CSMD2 missense variants and associated traits. **A.** Radar plot showing the percentage of individuals with biallelic *CSMD2* missense variants presenting each clinical feature. For each symptom, the percentage was calculated relative to the number of individuals for whom that specific information was available (see **Table S1** for details). **B.** Swiss-Pdb Viewer CSMD2 structure in ribbon representation color coded according to its domain architecture. The residues mutated in the affected individuals carrying biallelic missense variants are highlighted in white. **C.** Swiss-Pdb Viewer CSMD2 structure in ribbon representation color coded according to its domain architecture. The residues mutated in the affected individuals carrying *de novo* missense variants are highlighted in white.

ASD (autism spectrum disorder) (5/8), delayed speech and language development (5/8) and/or epilepsy-like symptoms (5/9), i.e. either generalized bilateral epileptiform activity, seizure-like activity or EEG alterations with large theta rhythm in the right posterior leads, in wakefulness and in sleep. Rarer symptoms present in the affected individuals, such as ADHD, facial dysmorphism and ataxia/spasticity, are summarized in **Figure 1A** and detailed in **Table S1**. The identified variants affect residues generally well conserved through evolution (**Figure S4**), between human CSMDs paralogs (**Figures S1-S2**) and within CUB and Sushi domains-containing proteins (**Figure S5**). While the eleven identified CSMD2 biallelic missense variants are spread along the encoded protein, 3D modelling of the predicted mutant proteins showed that a large fraction of the corresponding mutated residues are structurally close and forming two clusters. The first one with G85, A98 and R103 positioned in CUB1 and the second comprising R1609 (Sushi9), T1680 and Q1765 in CUB10, K2175 (Sushi12), T2190 (CUB13), R2445 (CUB14), and R2635 (Sushi16) located in the core CUB-Sushi region (**Figure 1B**). These missense variants are predicted to impair CSMD2 function as shown in **Figure S6** and detailed in **Table S3**, for example: (i) G85D introduces a negative charge that could affect molecular interactions; (ii) R1609W will impede both a hydrogen bond with G1611 and a salt bridge with D1600; (iii) Q1765R potentially disturbs the CUB10-CUB14 interaction by introducing a positive charge; (vi) R2445H hinders the formation of a salt bridge with E2410 and would clash with surrounding sidechains such as Y2443.

We assessed the effect of eight of the nine identified biallelic NDD-associated missense variants on CSMD2 localization by transfecting HEK293T cells with plasmids encoding N-terminal FLAG-tagged mouse Csmd2 WT and mutated isoforms. While we found no significant differences in the fraction of CSMD2 protein localized at the membrane between the WT and the mutated T2190I (p=1.0) or R2445H (p=0.087) proteins, we observe significantly less CSMD2 G85D (p=8.1e-11), R1609W (p=1.1e-11), Q1765R (p=8.8e-12), K2175M (p=3.0.e-14), R2635C (p=4.7e-8) and L3532R (p=5.5e-14) containing isoforms at the plasma membrane suggesting that some of the missense variants identified in the affected individuals present an impaired function through mislocalization and/or cytoplasmic aggregation (**Figure 2A-2C**).

**Figure 2.**
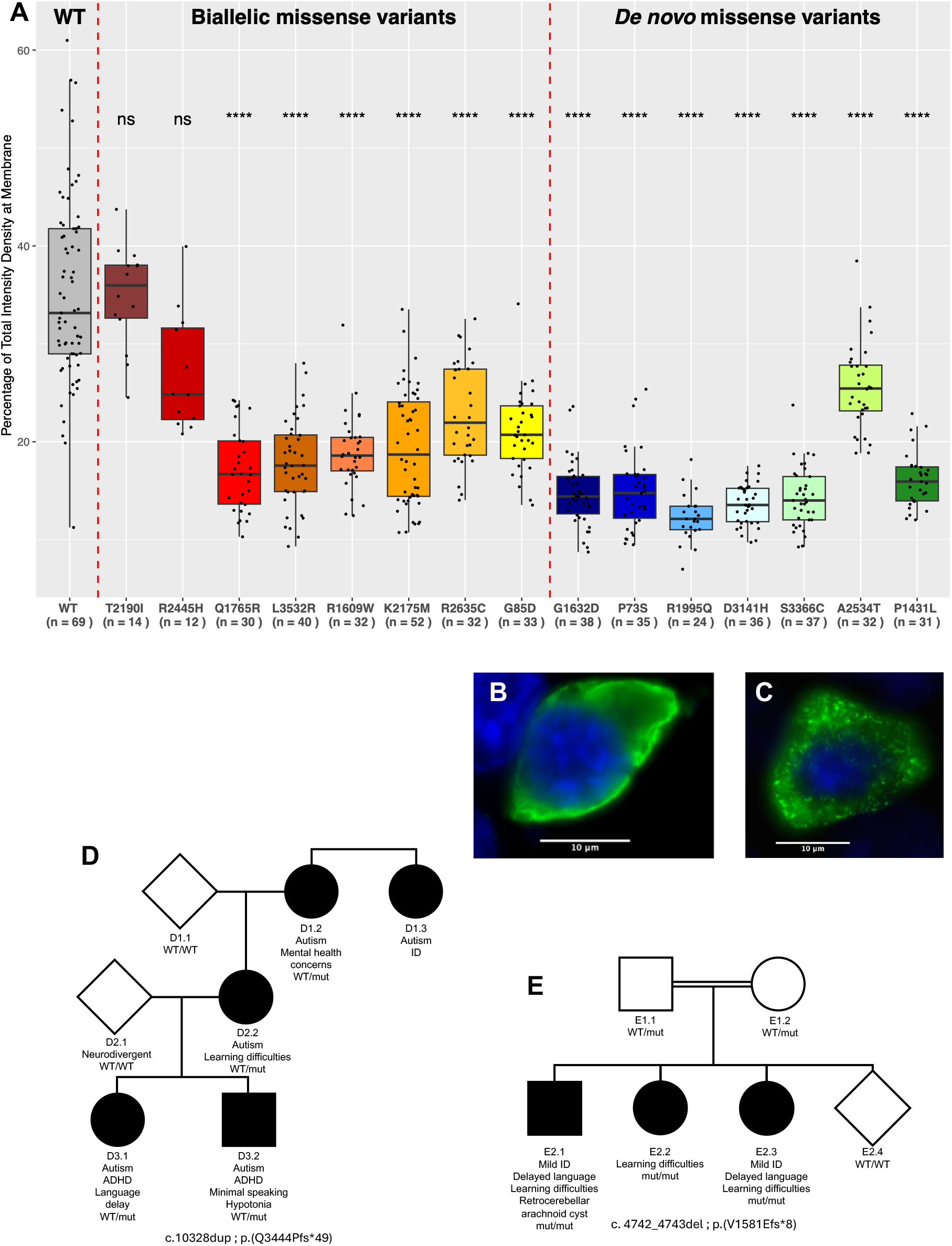
CSMD2 missense variants effect on cellular localization and pedigrees of families with truncating variants. **A.** Boxplot percentage of CSMD2 protein located at the membrane for each genotype. Performed in 3 independent batches, with reproducible data and no batch effect. The number of assessed cells is indicated on the x-axis. **B.** Image of a HEK293T cell transfected with the WT CSMD2 plasmid, after an immunolabelling, showing that the WT protein is mostly localized at the membrane. **C.** Image of a HEK293T cell transfected with the plasmid Q1765R, after an immunolabelling, showing that the Q1765R mutated protein presents a cytoplasmic aggregation. **D.** Pedigree of the family segregating the CSMD2 p.(Q3444Pfs*49) frameshift variant **E.** Pedigree of the family segregating the CSMD2 p.(V1581Efs*8) frameshift variant.

GnomAD v4.1.0^50,51^ population metrics indicate that *CSMD2* is under constraint for both truncation (pLOEUF=0.364; pLI=1) and missense variants (Z_missense_=5.48) suggesting that haploinsufficiency too, could be associated with diseases. This hypothesis is supported by a previously reported individual with ASD carrying a *de novo* (p.(G9AfsTer303) truncating variant^52^ (**Table S4**). We further identified a three-generation family with four affected individuals segregating a p.(Q3444PfsTer49) *CSMD2* frameshift. Affected family members presented with ASD and ADHD sometimes associated with ID and epilepsy (**Figure 2D, Table S4**). Suggestive of reduced penetrance, we also identified a family with consanguineous parents and three homozygous offsprings for the p.(V1581Efs*8) truncation variant (**Figure 2E**). Affected siblings presented with mild ID, delayed language development and learning difficulties.

We concomitantly identified four affected individuals with heterozygous *de novo* predicted-to-be deleterious *CSMD2* missense variants (P73S, D1227E, G1632D and Y2061C) (**Table S5**). Data supporting an association of such rare de novo *CSMD2* missense variants with pathologies have been reported in: (i) four affected individuals respectively affected by ASD (p.(R1995Q)), schizophrenia (p.(D2110G on alternative isoform NM_052896; not included in our analyses)), ID (p.(D3141H)) and epilepsy (p.(R155C))^53–56^; (ii) seven individuals (R133H, E1083K, P1431L, V1670I, P2347L, A2534T and S3366C) identified in a developmental disorders (DD) cohort of 31,058 parent-offspring trios compared to five synonymous *de novo* variants^57^ (**Table S5**). The predicted deleteriousness and frequencies of these *de novo* variants are detailed in **Table S2**. Like the biallelic variants, the *de novo* mutated residues cluster either in CUB1 (P73, R133 and R155) or in the core region spanning from Sushi 8 to CUB10 region, P1431 (Sushi8), G1632 (Sushi9), V1670 (CUB10), R1995 (Sushi11), Y2061 (CUB12) and P2347 (Sushi13) (**Figure 1B-1C)**. To evaluate function, we 3D modeled the encoded mutated protein (**Table S3**) and assessed the localization of these CSMD2 isoforms (**Figure 2A)**. We evaluated seven of the fourteen identified *de novo* missense variants mutated proteins by immunofluorescence: beside the two variants described here, they were selected to be representative of the variance in predicted deleteriousness (**Table S2**). A significantly lower fraction of P73S (p = 1.2e-13), P1431L (p = 1.8e-12), G1632D (p = 1.5e-14), R1995Q (p = 1.1e-10), A2534T (p = 3.4e-6), D3141H (p = 3.3e-14) and S3366C (p = 2.3e-14) isoforms was present at the cell membrane compared to WT (**Figure 2A)**.

A parallel analysis of the 84 and 10,390 UKBB participants, who carry rare *CSMD2* pLOF and missense variants, respectively, further supports both a haploinsufficiency model and causativeness of the *de novo* missense variants. While none of our tests survived Bonferroni correction, we identified three nominally significant (α = 0.05) associations: missense medium-confidence variants with ADHD (OR = 3.42, p = 0.019), missense high-confidence variants with childhood autism (OR = 4.13, p = 0.040), and pLoF variants with neuroticism (t = −2.01, p = 0.045).

### Zebrafish knock-down crispant larvae

The one-to-one ortholog of *CSMD2* in zebrafish, *csmd2*, is expressed in both neural and glial lineages between 5-and 120-hours post-fertilization (hpf), making zebrafish a suitable model to study its function during early neurodevelopment. We ablated *csmd2* using CRISPR-Cas9 genome editing. To bolster efficacy and ensure that F0 crispant larvae are hit on both alleles we used a combination of four different single guide RNA (sgRNA) targeting exon 4, 19, 24 and 56. These sgRNAs presented an individual efficiency of 100%, 80%, 100% and 67%, respectively. The crispant larvae presented a smaller head (p=0.00018) (**Figure S7**), and an overall shorter body (p=3.5e-05) (**Figure S8**). We also identified a statistically significant decrease in swimming velocity, both in the light (p=0.0097) and in the dark (p=0.00025) (**Figure 3A**), a decrease often linked to neuromotor deficits. This observation was further supported upon analysis of the motoneurons, as immunolabelling of acetylated tubulin showed a statistically significant increase of motoneurons with branching defects in the crispant larvae (p=2.3e-11) (**Figure 3B-3F**). Following the observation of the impact of the *csmd2* knockdown on the peripheral nervous system, we sought to determine whether the central nervous system was similarly impacted. First, we evaluated whether the cerebellum, which is involved in voluntary movements and motor learning, such as swimming, and in cognitive function, was similarly histologically altered by ablation of *csmd2*. The cerebellum area was significantly decreased in crispant larvae (p=7.6e-08) (**Figure 3G-3H**). Next, we counted and evaluated the size of the cerebellar Purkinje neurons. Following immunolabelling with an anti-parvalbumin 7 antibody, we found that while the number of Purkinje cells was similar in mock and crispant larvae (**Figure 3I**), their size was significantly reduced in crispant larvae (p<2.2e-16) (**Figure 3J**).

**Figure 3.**
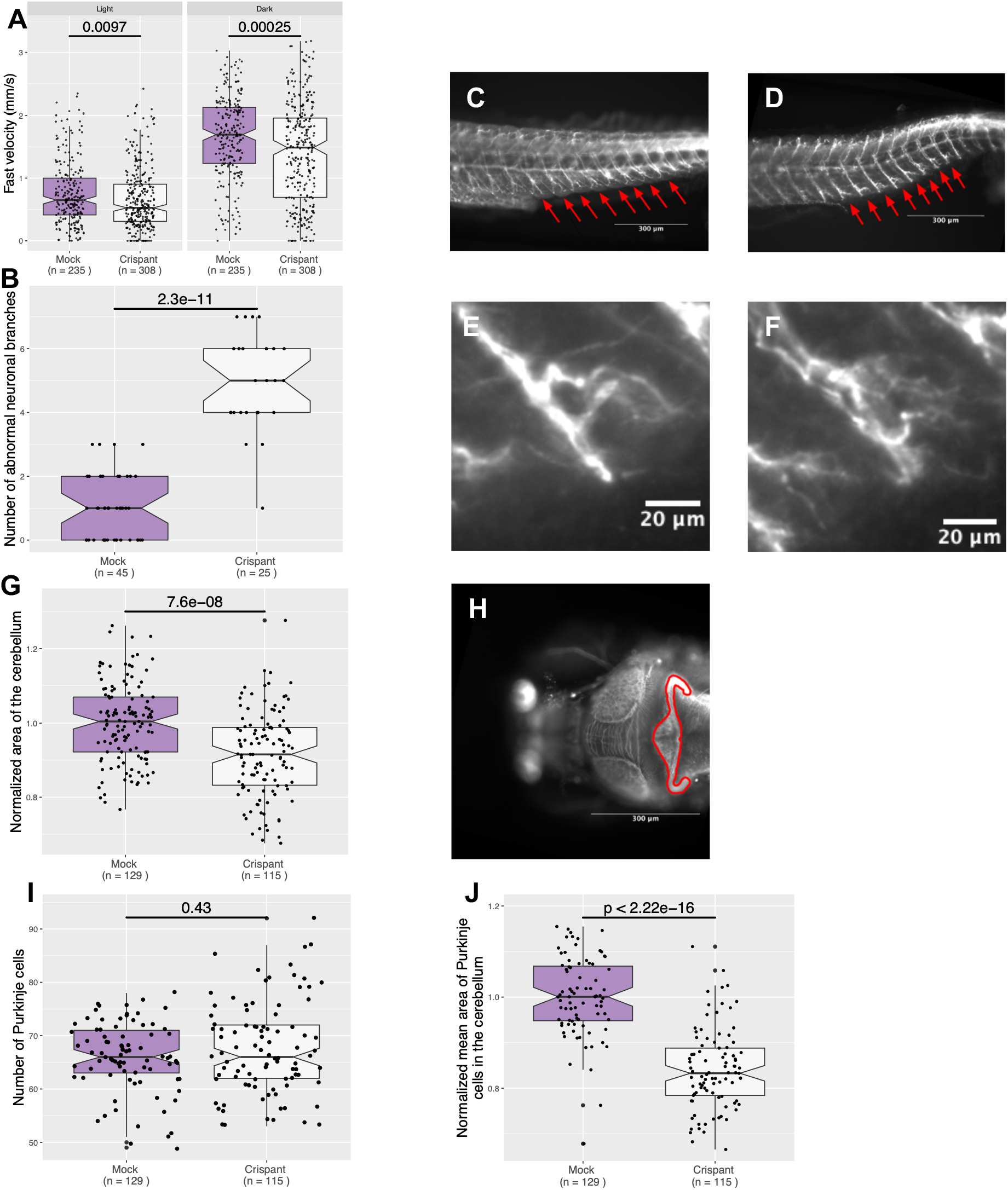
Effects of *csmd2* knockdown on velocity, peripheral neurons and the cerebellum. **A.** Fast velocity in mm/s in the light and in the dark, the movements of the larvae were measured using a Zebrabox apparatus, at 5 dpf. Fast velocity was calculated as detailed in Methods. **B.** Number of abnormal peripheral neuronal branches out of 8 in the tail of the larvae after an immunolabelling directed against acetylated tubulin at 4 dpf. **C.** Picture of the tail of a mock-injected larva after an immunolabelling directed against acetylated tubulin at 4 dpf. The red arrows highlight neuronal branches. **D.** Picture of the tail of a crispant larva after an immunolabelling directed against acetylated tubulin at 4 dpf. The red arrows highlight neuronal branches. **E-F.** Close-up pictures of abnormal neuronal branches from **D. G.** Normalized area of the cerebellum in mock-injected and crispant larvae after an immunolabelling directed against the acetylated tubulin at 4 dfp. **H.** Picture of the head of a mock-injected larva after an immunolabelling directed against acetylated tubulin at 4 dfp. The red line surrounds the area of the cerebellum. **I.** Number of Purkinje’s cell in half the cerebellum of mock-injected and crispant larvae after an immunolabelling directed against parvalbumin 7 at 4 dfp. **J.** Normalized mean area of Purkinje cells in the cerebellum of mock-injected and crispant larvae after immunolabelling directed against parvalbumin 7 at 4 dpf. The mean area was calculated from 10 individually measured cells per larva. For the graphs A, B, G, I and K, values below the 1st or above the 99th percentile were excluded as outliers prior to analysis. Boxes show the median and interquartile range, with notches indicating the 95% confidence interval around the median; whiskers represent the [Q1 - 1,5×IQR; Q3 + 1,5×IQR] interval. Statistical significance between mock-injected and crispant groups was assessed using a two-sided Wilcoxon rank-sum test. Normalization methods are detailed in Methods.

We then measured the minimum distance between the two optic tecta of the crispant larvae. This distance was significantly increased in crispant larvae (p=3.7e-06), the tecta being further apart from one another could be due to them being smaller when csmd2 expression is ablated (**Figure 4A-4B**). Second, the measurement of the area of one optic tectum per larvae, showed a significant decrease in crispant larvae compared to mock larvae (p=4.8e-10), confirming that the loss of *csmd2* leads to microcephaly (**Figure 4C-D**).

**Figure 4.**
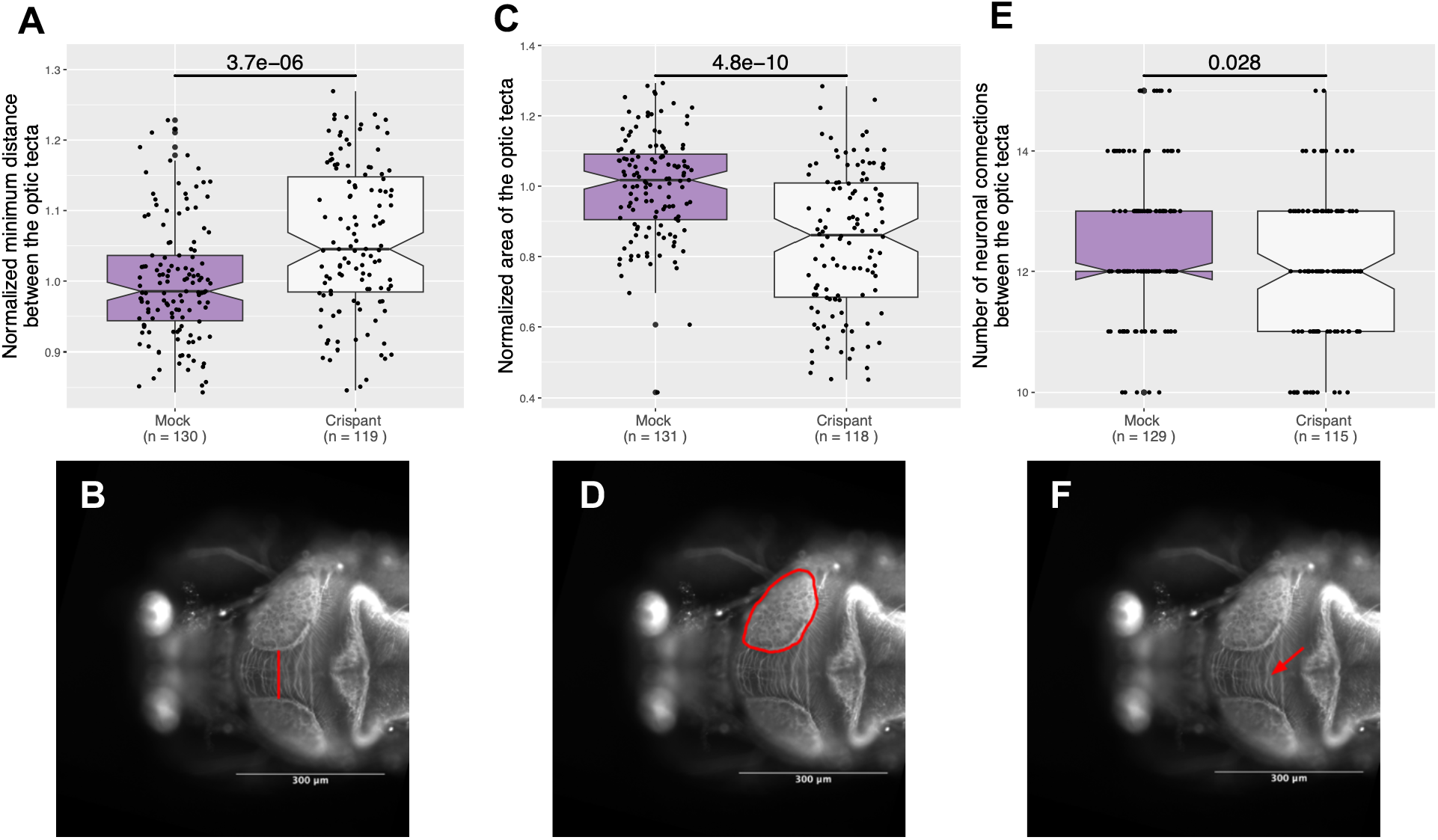
Effects of *csmd2* knockdown on the brain. **A.** Normalized minimum distance measured between the two optic tecta in mock-injected and crispant larvae after an immunolabelling directed against acetylated tubulin at 4 dpf. **B.** Picture of the head of a mock-injected larva after an immunolabelling directed against acetylated tubulin at 4 dpf, the red line highlights the minimum distance between the optic tecta. **C.** Normalized area of the optic tecta of mock-injected and crispant larvae after an immunolabelling directed against acetylated tubulin at 4 dpf. **D.** Picture of the head of a mock-injected larva after an immunolabelling directed against acetylated tubulin at 4 dpf. The red line surrounds the area of the optic tecta. **E.** Number axonal tracts counted between the two optic tecta in mock-injected and crispant larvae after an immunolabelling directed against acetylated tubulin at 4 dpf. **F.** Picture of the head of a mock-injected larva after an immunolabelling directed against acetylated tubulin at 4 dpf. Tthe red arrow highlights axonal tracts between the optic tecta. For the graphs A, C and E, values below the 1st or above the 99th percentile were excluded as outliers prior to analysis. Boxes show the median and interquartile range, with notches indicating the 95% confidence interval around the median; whiskers represent the [Q1 - 1,5×IQR; Q3 + 1,5×IQR] interval. Statistical significance between mock-injected and crispant groups was assessed using a two-sided Wilcoxon rank-sum test. Normalization methods are detailed in Methods.

To further assess the impact of *csmd2* loss on the development of the central nervous system, we compared the number of inter-tecta axonal tracts, and we found significantly less axonal tracts in crispant larvae compared to mock larvae (p=0.028) (**Figure 4E-4F**).

## DISCUSSION

We present evidence suggesting that multiple *CSMD2-*related inheritance patterns are associated with NDD akin to recent examples^58–60^ and in line with the notion that human autosomal recessive disorders have widespread effects in heterozygotes^61^. Firstly, we identified nine individuals from eight unrelated families with biallelic variants in *CSMD2*. They present with a NDD characterized by ID, brain anomalies, epilepsy-like seizures, ADHD and ASD. Secondly, we discovered a family segregating a likely dominant *CSMD2* truncation variant with ASD/ADHD and a consanguineous family with siblings homozygous for a *CSMD2* truncation variant presenting with mild ID. Importantly, we can neither rule out a mild phenotype in the heterozygous parents of the second family, who were not thoroughly phenotypically examined, nor the possibility of alternative splicing mitigating the effect of the second truncation variant as we do not have access to the suitable tissue samples. Thirdly, we concomitantly show that biobank volunteers carrying *CSMD2* truncation and rare missense variants are nominally enriched for neuroticism, ADHD and ASD. Accordingly, we describe fourteen affected individuals carrying predicted-to-be deleterious *de novo* missense variants. The CSMD2 residues found to be mutated in the three groups of affected individuals: “biallelic NDD”, “biallelic epilepsy” described in reference^31^ and *de novo* heterozygous present similar level of conservation through evolution, i.e. in orthologs (**Figure S1, Figure S2)** and in human paralogs (**Figure S4**). Our study is not without limitations, for example, the phenotypes of the affected individuals are not fully overlapping. These pathologies are likewise not particularly rare or specific, which in turn does not allow ruling out spurious association between rare predicted-to-be deleterious variants and the observed traits. To mitigate this possibility, we assessed the cellular localization in an ectopic expression system (which has its own limits) of the encoded mutated CSMD2 isoforms carrying either a recessive or a *de novo* missense variant. For 87% of the tested variants (13 out of 15), we found that a significant fraction of the mutated isoform was not positioned at the membrane but aggregated in the cytoplasm. Overall, the *de novo* missense heterozygous variants seem to have a more severe impact on protein localization and cytoplasmic aggregation than biallelic missense variants consistent with their increased disease liability (**Figure 2A**). Interestingly, the three isoforms that are less perturbed in their localization, i.e. T2190I (biallelic), R2445H (biallelic) and A2534T (*de novo*) (**Figure 2A),** are all positioned within the CUB13 to Sushi15 core region (**Figure 1B-C**). The evaluation of the pathogenicity of the missense variants via zebrafish rescue experiments with *Csmd2* WT and mutated mRNAs as described^34,58,62^, was not successful since we were unable (after repeated attempts) to produce full-length RNA *in vitro* possibly due to the large size and/or high GC content of the corresponding cDNA (>10kb long and 57% GC). Compounding data for each of the twenty-five missense variants reported here, i.e. 3D modeling, encoded protein localization, residue conservation and prediction of deleteriousness, support causativeness for many of these variants (**Figure 5** and **Table S2, Table S3**). Previously published *de novo* missense D1227E^63^ and A2534T^57^ and compound heterozygous variants A98T and T1680S^31^ have the weakest support and should be considered VUS at this stage. While the compound heterozygous variants K2175M and R2445H described in this report have likewise insufficient support for pathogenicity, they are paired with R2635C and Q1765R, respectively, two missense variants with stronger support (**Figure 5** and **Table S2, Table S3**). Our *csmd2* crispant knockout zebrafish larvae further showcase the role of *CSMD2* in the development of both the central and peripheral nervous system. Its impact on the development of both the brain and the cerebellum could be linked with the cognitive deficiency observed in our affected individuals.

**Figure 5.**
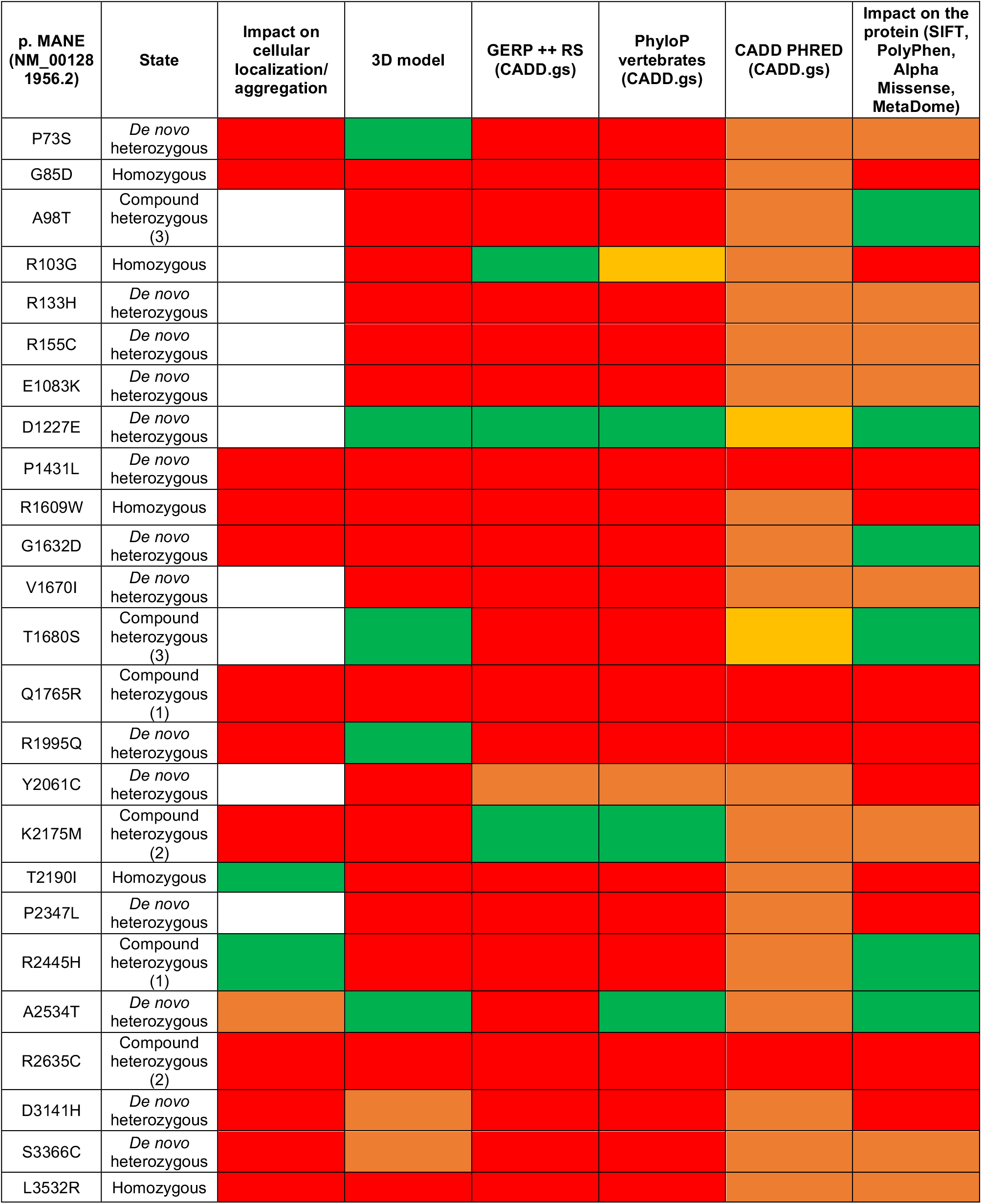
Compound effect of *CSMD2* missense variants. Heatmap compiling the inheritance state of the variant, impact on cellular localization/aggregation (experimentally determined, see Figure 2A; red: altered localization/aggregation observed, white: not tested; Green: no alteration observed), 3D modelling impact prediction (red: deleterious; orange: uncertain; green: benign), GERP++ RS (from CADD.gs, green: RS<3 low conservation; orange: 3≤RS<4.4 medium conservation; red: RS≥4.4 high conservation), PhyloP vertebrates (from CADD.gs, red: >3 extreme conservation cutoff; orange: 2.27–3 high confidence cutoff; pale orange: 1.6–2.27 standard cutoff; green: <1.6 below cutoff), CADD PHRED (from CADD.gs, red: ≥30, top 0.1% most deleterious substitutions genome-wide; orange: ≥20, top 1% most deleterious; pale orange: ≥10, top 10% most deleterious), and Impact on the protein (SIFT, PolyPhen, AlphaMissense, MetaDome combined per the following rule, green: 4 green or 3 green + 1 orange or 3 green + 1 red; Orange: 2 green + 2 red or 2 red + 1 orange + 1 green or 2 orange + 1 green + 1 red; Red: 4 red or 3 red + 1 green or 3 red + 1 orange. With SIFT (Red: < 0.05 deleterious; Green: ≥ 0.05 tolerated), PolyPhen (Green: < 0.15 benign; Orange: 0.15–0.85 possibly damaging; Red: > 0.85 probably damaging), AlphaMissense (Green: < 0.34 likely benign; Orange: 0.34–0.564 ambiguous; Red: > 0.564 likely pathogenic), MetaDome tolerance score (Red: < 0.20 highly intolerant, 0.20–0.49 intolerant and 0.50–0.69 slightly intolerant; Orange: 0.70–0.99 neutral; Green: 1.00–1.29 tolerant and ≥ 1.30 (highly tolerant). The individuals scores are detailed in **Table S2**.

While one of the six individuals affected with focal epilepsy carrying *CSMD2* bi-allelic variants described in reference^31^ was also diagnosed with mild ID, the remaining five individuals had a normal development compatible with the notions of pleiotropy and variable expressivity. Parallels can be drawn for the paralogous *CSMD1 and CSMD3* for which multiple inheritance mechanisms and pleiotropy might also be at play. GnomAD v4.1.0 population metrics suggest that both *CSMD1 and CSMD3* could be haploinsufficient with pLOEUF=0.586 and =0.464, respectively^50,51^. Biallelic variants in *CSMD1* were recently associated with ID, cortical malformations, epileptic encephalopathy and general epilepsies^20,32^. Consistent with this association, we identified a consanguineous Pakistani family with two affected siblings homozygous for a stop-gain variant in *CSMD1*, MANE NM_033225:c.5140C>T:p.R1714X and matching traits within our cohort of exome-sequenced multiplex NDD consanguineous families (**Figure S3B, Table S6**). *CSMD1 de novo* missense or truncation variants were also identified in nineteen and two individuals presenting with DD and epilepsy, respectively^32,57^. Likewise, autosomal recessive and dominant inheritance were suggested for *CSMD3*-associated diseases^31^, for example four *de novo* missense or truncation variants in a trio DD cohort^57^. Homozygous ablation of murine *Csmd1* and *Csmd3* were associated with behavioral/neurological, growth and metabolic impairments^15,28,64,65^. These results suggest that the *CSMD* paralogs are not redundant, a hypothesis further supported by their only-partially overlapping expression patterns through development.

In conclusion and notwithstanding the above-cited limitations, we provide evidence for involvement of rare likely pathogenic *CSMD2* variants in NDD with both autosomal recessive and dominant inheritance. Our results compounded with the previous GWAS associations between neurological traits and common *CSMD2* variants^14–19^ further strengthen the evidence linking *CSMDs* with neurodevelopment.

## Data Availability

All data produced in the present study are available upon reasonable request to the authors

## ACKNOWLEDGMENTS

We are grateful to Dr Santos J. Franco, University of Colorado School of Medicine for providing materials, to Jacques S. Beckmann and Federico Santoni for comments on the manuscript and to GeneDx for their correspondence regarding this work. This work was supported by grants from the Swiss National Science Foundation (31003A_182632 to AR; IZSTZ0_216615 and 3200-0-242970 to AR and MA), the Blackswan Foundation (AR), the ChildCare Foundation (SEA), the Department of Computational Biology (ZK) and the Center for Integrative Genomics (AR) from the University of Lausanne, the Agence Nationale de la Recherche (ANR-22-CE12-0011 to CG), the Fondation de France (WB-2022-45868 to CG), the French National Research Agency (ANR) through the Programme d’Investissement d’Avenir under contract ANR-10-LABX-0030-INRT grant under the frame programme Investissement d’Avenir ANR-10-IDEX-0002-02 and ITMO Cancer of Aviesan within the framework of the 2021-2030 Cancer Control Strategy on funds administered by Inserm. This work of the Interdisciplinary Thematic Institute IMCBio+, as part of the ITI 2021-2028 program of the University of Strasbourg, CNRS and Inserm, was supported by IdEx Unistra (ANR-10-IDEX-0002), and by SFRI-STRAT’US project (ANR-20-SFRI-0012) and EUR IMCBio (ANR-17-EURE-0023) under the framework of the France 2030 Progr. CG is a permanent INSERM investigator, MVL is a doctoral fellow supported by EUR IMCBio funds and Fondation de France (WB-2022-45868) and FN is a member of the European Reference Network for Rare Neurological Diseases (Project ID No 739510). Sequencing and analysis of some samples were provided by the Broad Institute of MIT and Harvard Center for Mendelian Genomics (Broad CMG) and was funded by the National Human Genome Research Institute, the National Eye Institute, and the National Heart, Lung and Blood Institute grant UM1HG008900. The content is solely the responsibility of the authors and does not necessarily represent the official views of the National Institutes of Health. UK Biobank has approval from the NorthWest Multicentre Research Ethics Committee as a Research Tissue Bank. All participants signed a broad informed consent form. Data were accessed under application 16389. The funders had no role in study design, data collection and analysis, decision to publish, or preparation of the manuscript.

## AUTHORS’ CONTRIBUTIONS

CPP, FM, HD, GZ, GSC, LT, FN, SMA, CL, JGG, IS, LF, MSZ, LB, EBB, PK, NF, VL, CN, TS, CVD, ZI, MT, TZ, SKJ, AF, MU, ZA, TS, SF, SGF, AT, SS, FJ, MB, FH, EOH, LP, ASJ, MET, QW, LK, TK, SEA, and MA collected clinical information and DNAs, sequenced and analyzed exomes and/or genomes. NG and GA 3D-modeled missense variants. CPP, MVL, FM, SB, JC, and CG engineered and phenotyped animal models. CC and ZK analyzed UK Biobank data. CPP and AR wrote the manuscript. All other authors read and approved the manuscript. AR conceived and supervised the study.

---

**Figure S1** Comparison of the CSMD1, CSMD2 and CSMD3 protein sequences. The residues conserved through two of the three paralogs are in light blue, while the ones conserved in all three of the paralogs are in dark blue. The biallelic mutated CSMD2 residues inherited in the affected individuals described in this report are highlighted in red, while the *de novo* mutations are highlighted in green. The residues reported mutated in references ^31^ and ^32^ and associated with epilepsy are in pink. The mutated CSMD1 residues identified in NDD individuals and reported in references ^20^ and ^33^ are highlighted in orange. This alignment was performed on the UniProt website1 using the MANE transcript of each protein (Homo_sapiens_ENSP00000489225_1 for CSMD1, Homo_sapiens_ENSP00000362479_4 for CSMD2 and Homo_sapiens_ENSP00000297405_5 for CSMD3).

**Figure S2** Phylogenetic tree based on the alignment of the CSMD2 protein sequences. This alignment was performed on the UniProt website1.

**Figure S3 Pedigrees. A.** Pedigree of the family segregating the CSMD2 p.(T2190I) missense variant. Phenotypes of the affected individuals are detailed in **Table S1**. **B.** Pedigree of the family segregating the CSMD1 p.(R1714X) stop-gain variant. Phenotypes of the affected individuals are detailed in **Table S6**.

**Figure S4** Comparison of the CSMD2 protein sequences between ten vertebrate species. The darker the background of a residue, the more it is conserved. The biallelic mutated CSMD2 residues inherited in the affected individuals described in this report are highlighted in red, the *de novo* mutations are highlighted in green. The residues reported mutated in reference ^31^ and linked to epilepsy are in pink. This alignment was performed on the UniProt website1.

**Figure S5 Metadome diagrams.** Scheme showing the tolerance landscape for CSMD1 (**A**), CSMD2 (**B**) and CSMD3 (**C**) across species and across homologous human protein domains, respectively. The biallelic mutated CSMD2 residues inherited in the affected individuals described in this report are highlighted in red, the de novo mutations in green, residues reported mutated in references ^31^ and ^32^ and linked to epilepsy are in pink. The CSMD1 residues identified in NDD individuals and reported in references ^20^ and ^33^ are highlighted in orange.

**Figure S6 Swiss-Pdb Viewer CSMD2 structure in ribbon representation. A.** Close-up view of R1609 of human CSMD2 protein. **B.** W1609 replacing the WT Arginine. **C.** Close −up view of Q1765 of human CSMD2 protein (Q1725 on AF-Q7Z408_v2 model). **D.** R1765 (R1725 on AF-Q7Z408_v2 model) replacing the WT Glutamine. **E.** Close-up view of R2445 of human CSMD2 protein. **F.** H2445 replacing the WT Arginine. Detailed descriptions are given in the text and **Table S3.**

**Figure S7 Effects of *csmd2* knockdown on zebrafish head size A.** Picture of the head of a depigmented mock-injected larva. The red line shows where the head size was measured (magnification 6.3x). **B.** Head size at the level of the pupils in mock-injected and crispant larvae. Values below the 1st or above the 99th percentile were excluded as outliers prior to analysis. Boxes show the median and interquartile range, with notches indicating the 95% confidence interval around the median; whiskers represent the [Q1 −1,5×IQR; Q3 + 1,5×IQR] interval. Statistical significance between mock-injected and crispant groups was assessed using a two-sided Wilcoxon rank-sum test. Normalization was performed as detailed in Methods.

**Figure S8 Effects of *csmd2* knockdown on zebrafish body length A.** Picture of a depigmented mock-injected larva. The red segmented line shows body length measurement (magnification 2x). **B.** Body length, from the mouth to the tip of the tail in mock-injected and crispant larvae. Values below the 1st or above the 99th percentile were excluded as outliers prior to analysis. Boxes show the median and interquartile range, with notches indicating the 95% confidence interval around the median; whiskers represent the [Q1 −1,5×IQR; Q3 + 1,5×IQR] interval. Statistical significance between Mock and crispant groups was assessed using a two-sided Wilcoxon rank-sum test. Normalization was performed as detailed in Methods.

**Table S1 Genotype and phenotype of individuals with *CSMD2* biallelic missense variants**

**Table S2 Predicted deleteriousness of *CSMD2* variants**

**Table S3 3D model: position and predictive effects of the mutations observed in CSMD2**

**Table S4 Genotype and phenotype of individuals with CSMD2 truncating variants**

**Table S5 Genotype and phenotype of individuals with CSMD2 *de novo* missense variants**

**Table S6 Genotype and phenotype of *CSMD1* homozygous stop-gain siblings**

