## Supplementary material for "Haploinsufficiency, *de novo* and biallelic missense variants in *CSMD2* are associated with neurodevelopmental disorders": Figure S1

```
CSMD1-217 -----------------------------------------------MTAWRRFQ-- 8
CSMD2-204 MP----------RSRGRELG-----RCGCPAGRARGETGISALVPGAGSRWGRPPPP 42
CSMD3-201 MKGIRKGESRAKESKPWEPGKRRCAKCGRLDFILMKKMGI----KSGFTFWN----- 48

CSMD1-217 ---SLLLLLGLLVLCARLLTAAKGQNCGGLVQGPNGTIESPGFPHGYPNYANCTWII 62
CSMD2-204 TPPPLLLLLGCGL---LSVSAAAGQNCTFQLHGPNGTVESPGFPYGYPNYANCTWTI 96
CSMD3-201 ----------LVFLLTVSCVKGFIYTCGGTLKGLNGTIESPGFPYGYPNGANCTWVI 95

CSMD1-217 ITGERNRIQLSFHTFALEEDFDILSVYDGQPQQGNLKVRLSGFQLPSSIVSTGSILT 119
CSMD2-204 TAEEQHRIQLVFQSFALEEDFDVLSVFDGPPQPENLRTRLTGFQLPATIVSAATTLS 153
CSMD3-201 IAEERNRIQIVFQSFALEEEYDYLSLYDGHPHPTNFRTRLTGFHLPPPVTSTKSVFS 152

CSMD1-217 LWFTTDFAVSAQGFKALYEVLPSHTCGNPGEILKGVLHGTRFNIGDKIRYSCLPGYI 176
CSMD2-204 LRLISDYAVSAQGFHATYEVLPSHTCGNPGRLPNGIQQGSTFNLGDKVRYSCNLGFF 210
CSMD3-201 LRLTSDFAVSAHGFKVYYEELQSSSCGNPGVPPKGVLYGTRFDVGDKIRYSCVTGYI 209

CSMD1-217 LEGHAILTCIVSPGNGASWDFPAPFCRAEGACGGTLRGTSSSISSPHFPSEYENNAD 233
CSMD2-204 LEGHAVLTCHAGSENSATWDFPLPSCRADDACGGTLRGQSGIISSPHFPSEYHNNAD 267
CSMD3-201 LDGHPQLTCIANSVNTASWDFPVPICRAEDACGGTMRGSSGIISSPSFPNEYHNNAD 266

CSMD1-217 CTWTILAEPGDTIALVFTDFQLEEGYDFLEISGTEAPSIWLTGMNLPSPVISSKNWL 290
CSMD2-204 CTWTILAELGDTIALVFIDFQLEDGYDFLEVTGTEGSSLWFTGASLPAPVISSKNWL 324
CSMD3-201 CTWTIVAEPGDTISLIFTDFQMEEKYDYLEIEGSEPPTIWLSGMNIPPPIISNKNWL 323

CSMD1-217 RLHFTSDSNHRRKGFNAQFQ------------------------------------- 310
CSMD2-204 RLHFTSDGNHRQRGFSAQYQ------------------------------------- 344
CSMD3-201 RLHFVTDSNHRYRGFSAPYQGSSTLTHTTSTGELEEHNRTTTGAIAVASTPADVTVS 380

CSMD1-217 --------------------------------------------------------- 310
CSMD2-204 --------------------------------------------------------- 344
CSMD3-201 SVTAVTIHRLSEEQRVQVTSLRNSGLDPNTSKDGLSPHPADTQSTRRRPRHAEQIER 437

CSMD1-217 ----------VKKAIELKSRGVKMLPSKDGSHKNSVLSQGGVALVSDMCPDPGIPEN 357
CSMD2-204 ----------VKKQIELKSRGVKLMPSKDNSQKTSVLTQVGVSQGHNMCPDPGIPER 391
CSMD3-201 TKELAVVTHRVKKAIDFKSRGFKLFPGKDNSNKFSILNEGGIKTASNLCPDPGEPEN 494

CSMD1-217 GRRAGSDFRVGANVQFSCEDNYVLQGSKSITCQRVTETLAAWSDHRPICRARTCGSN 414
CSMD2-204 GKRLGSDFRLGSSVQFTCNEGYDLQGSKRITCMKVSDMFAAWSDHRPVCRARMCDAH 448
CSMD3-201 GKRIGSDFSLGSTVQFSCDEDYVLQGAKSITCQRIAEVFAAWSDHRPVCKVKTCGSN 551

CSMD1-217 LRGPSGVITSPNYPVQYEDNAHCVWVITTTDPDKVIKLAFEEFELERGYDTLTVGDA 471
CSMD2-204 LRGPSGIITSPNFPIQYDNNAHCVWIITALNPSKVIKLAFEEFDLERGYDTLTVGDG 505
CSMD3-201 LQGPSGTFTSPNFPFQYDSNAQCVWVITAVNTNKVIQINFEEFDLEIGYDTLTIGDG 608

CSMD1-217 GKVGDTRSVLYVLTGSSVPDLIVSMSNQMWLHLQSDDSIGSPGFKAVYQEIEKGGCG 528
CSMD2-204 GQDGDQKTVLYILTGTSVPDLIVSTNHQMWLLFQTDGSGSSLGFKASYEEIEQGSCG 562
CSMD3-201 GEVGDPRTVLQVLTGSFVPDLIVSMSSQMWLHLQTDESVGSVGFKVNYKEIEKESCG 665

CSMD1-217 DPGIPAYGKRTGSSFLHGDTLTFECPAAFELVGERVITCQQNNQWSGNKPSCVFSCF 585
CSMD2-204 DPGIPAYGRREGSRFHHGDTLKFECQPAFELVGQKAITCQKNNQWSAKKPGCVFSCF 619
CSMD3-201 DPGTPLYGIREGDGFSNRDVLRFECQFGFELIGEKSIVCQENNQWSANIPICIFPCL 722

CSMD1-217 FNFTASSGIILSPNYPEEYGNNMNCVWLIISEPGSRIHLIFNDFDVEPQFDFLAVKD 642
CSMD2-204 FNFTSPSGVVLSPNYPEDYGNHLHCVWLILARPESRIHLAFNDIDVEPQFDFLVIKD 676
CSMD3-201 SNFTAPMGTVLSPDYPEGYGNNLNCIWTIISDPGSRIHLSFNDFDLESQFDFLAVKD 779

CSMD1-217 DGISDITVLGTFSGNEVPSQLASSGHIVRLEFQSDHSTTGRGFNITYTTFGQNECHD 699
CSMD2-204 GATAEAPVLGTFSGNQLPSSITSSGHVARLEFQTDHSTGKRGFNITFTTFRHNECPD 733
CSMD3-201 GDSPESPILGTFTGAEVPSHLTSNSHILRLEFQADHSMSGRGFNITYNTFGHNECPD 836

CSMD1-217 PGIPINGRRFGDRFLLGSSVSFHCDDGFVKTQGSESITCILQDGNVVWSSTVPRCEA 756
CSMD2-204 PGVPVNGKRFGDSLQLGSSISFLCDEGFLGTQGSETITCVLKEGSVVWNSAVLRCEA 790
CSMD3-201 PGIPINARRFGDNFQLGSSISVICEEGFIKTQGTETITCILMDGKVMWSGLIPKCGA 893

CSMD1-217 PCGGHLTASSGVILPPGWPGYYKDSLHCEWIIEAKPGHSIKITFDRFQTEVNYDTLE 813
CSMD2-204 PCGGHLTSPSGTILSPGWPGFYKDALSCAWVIEAQPGYPIKITFDRFKTEVNYDTLE 847
CSMD3-201 PCGGHFSAPSGVILSPGWPGYYKDSLNCEWVIEAEPGHSIKITFERFQTELNYDVLE 950

CSMD1-217 VRDGPASSSPLIGEYHGTQAPQFLISTGNFMYLLFTTDNSRSSIGFLIHYESVTLES 870
CSMD2-204 VRDGRTYSAPLIGVYHGTQVPQFLISTSNYLYLLFSTDKSHSDIGFQLRYETITLQS 904
CSMD3-201 VHDGPNLLSPLLGSYNGTQVPQFLFSSSNFIYLLFTTDNSRSNNGFKIHYESVTVNT 1007

CSMD1-217 DSCLDPGIPVNGHRHGGDFGIRSTVTFSCDPGYTLSDDEPLVCERNHQWNHALPSCD 927
CSMD2-204 DHCLDPGIPVNGQRHGNDFYVGALVTFSCDSGYTLSDGEPLECEPNFQWSRALPSCE 961
CSMD3-201 YSCLDPGIPVHGRRYGHDFSIGSTVSFSCDSGYRLSHEEPLLCEKNHWWSHPLPTCD 1064

CSMD1-217 ALCGGYIQGKSGTVLSPGFPDFYPNSLNCTWTIEVSHGKGVQMIFHTFHLESSHDYL 984
CSMD2-204 ALCGGFIQGSSGTILSPGFPDFYPNNLNCTWIIETSHGKGVFFTFHTFHLESGHDYL 1018
CSMD3-201 ALCGGDVRGPSGTILSPGYPEFYPNSLNCTWTVDVTHGKGVQFNFHTFHLEDHHDYL 1121

CSMD1-217 LITEDGSFSEPVARLTGSVLPHTIKAGLFGNFTAQLRFISDFSISYEGFNITFSEYD 1041
CSMD2-204 LITENGSFTQPLRQLTGSRLPAPISAGLYGNFTAQVRFISDFSMSYEGFNITFSEYD 1075
CSMD3-201 LITENGSFTQPLARLTGSDLPPTINAGLYGNFRAQLRFISDFSISYEGFNITFSEYN 1178
```

```
CSMD1-217 LEPCDDPGVPAFSRRIGFHFGVGDSLTFSCFLGYRLEGATKLTCLGGGRRVWSAPLP 1098
CSMD2-204 LEPCEEPEVPAYSIRKGLQFGVGDTLTFSCFPGYRLEGTARITCLGGRRRLWSSPLP 1132
CSMD3-201 LEPCEDPGIPQYGSRIGFNFGIGDTLTFSCSSGYRLEGTSEIICLGGGRRVWSAPLP 1235

CSMD1-217 RCVAECGASVKGNEGTLLSPNFPSNYDNNHECIYKIETEAGKGIHLRTRSFQLFEGD 1155
CSMD2-204 RCVAECGNSVTGTQGTLLSPNFPVNYNNNHECIYSIQTQPGKGIQLKARAFELSEGD 1189
CSMD3-201 RCVAECGASATNNEGILLSPNYPLNYENNHECIYSIQVQAGKGINISARTFHLAQGD 1292

CSMD1-217 TLKVYDGKDSSSRPLGTFTKNELLGLILNSTSNHLWLEFNTNGSDTDQGFQLTYTSF 1212
CSMD2-204 VLKVYDGNNNSARLLGVFSHSEMMGVTLNSTSSSLWLDFITDAENTSKGFELHFSSF 1246
CSMD3-201 VLKIYDGKDKTTHLLGAFTGASMRGLTLSSTSNQLWLEFNSDTEGTDEGFQLVYTSF 1349

CSMD1-217 DLVKCEDPGIPNYGYRIRDEGHFTDTVVLYSCNPGYAMHGSNTLTCLSGDRRVWDKP 1269
CSMD2-204 ELIKCEDPGTPKFGYKVHDEGHFAGSSVSFSCDPGYSLRGSEELLCLSGERRTWDRP 1303
CSMD3-201 ELSHCEDPGIPQFGYKISDQGHFAGSTIIYGCNPGYTLHGSSLLKCMTGERRAWDYP 1406

CSMD1-217 LPSCIAECGGQIHAATSGRILSPGYPAPYDNNLHCTWIIEADPGKTISLHFIVFDTE 1326
CSMD2-204 LPTCVAECGGTVRGEVSGQVLSPGYPAPYEHNLNCIWTIEAEAGCTIGLHFLVFDTE 1360
CSMD3-201 LPSCIAECGGRFKGESSGRILSPGYPFPYDNNLRCMWMIEVDPGNIVSLQFLAFDTE 1463

CSMD1-217 MAHDILKVWDGPVDSDILLKEWSGSALPEDIHSTFNSLTLQFDSDFFISKSGFSIQF 1383
CSMD2-204 EVHDVLRIWDGPVESGVLLKELSGPALPKDLHSTFNSVVLQFSTDFFTSKQGFAIQF 1417
CSMD3-201 ASHDILRVWDGPPENDMLLKEISGSLIPEGIHSTLNIVTIQFDTDFYISKSGFAIQF 1520

CSMD1-217 STSIAATCNDPGMPQNGTRYGDSREAGDTVTFQCDPGYQLQGQAKITCVQLNNRFFW 1440
CSMD2-204 SVSTATSCNDPGIPQNGSRSGDSWEAGDSTVFQCDPGYALQGSAEISCVKIENRFFW 1474
CSMD3-201 SSSVATACRDPGVPMNGTRNGDGREPGDTVVFQCDPGYELQGEERITCIQVENRYFW 1577

CSMD1-217 QPDPPTCIAACGGNLTGPAGVILSPNYPQPYPPGKECDWRVKVNPDFVIALIFKSFN 1497
CSMD2-204 QPSPPTCIAPCGGDLTGPSGVILSPNYPEPYPPGKECDWKVTVSPDYVIALVFNIFN 1531
CSMD3-201 QPSPPVCIAPCGGNLTGSSGFILSPNFPHPYPHSRDCDWTITVNADYVISLAFISFS 1634

CSMD1-217 MEPSYDFLHIYEGEDSNSPLIGSYQGSQAPERIESSGNSLFLAFRSDASVGLSGFAI 1554
CSMD2-204 LEPGYDFLHIYDGRDSLSPLIGSFYGSQLPGRIESSSNSLFLAFRSDASVSNAGFVI 1588
CSMD3-201 IEPNYDFLYIYDGPDSNSPLIGSFQDSKLPERIESSSNTMHLAFRSDGSVSYTGFHL 1691

CSMD1-217 EFKEKPREACFDPGNIMNGTRVGTDFKLGSTITYQCDSGYKILDPSSITCVIGADGK 1611
CSMD2-204 DYTENPRESCFDPGSIKNGTRVGSDLKLGSSVTYYCHGGYEVEGTSTLSCILGPDGK 1645
CSMD3-201 EYKAKLRESCFDPGNIMNGTRLGMDYKLGSTVTYYCDAGYVLQGYSTLTCIMGDDGR 1748

CSMD1-217 PSWDQVLPSCNAPCGGQYTGSEGVVLSPNYPHNYTAGQICLYSITVPKEFVVFGQFA 1668
CSMD2-204 PVWNNPRPVCTAPCGGQYVGSDGVVLSPNYPQNYTSGQICLYFVTVPKDYVVFGQFA 1702
CSMD3-201 PGWNRALPSCHAPCGSRSTGSEGTVLSPNYPKNYSVGHNCVYSIAVPKEFVVFGQFV 1805

CSMD1-217 YFQTALNDLAELFDGTHAQARLLSSLSGSHSGETLPLATSNQILLRFSAKSGASARG 1725
CSMD2-204 FFHTALNDVVEVHDGHSQHSRLLSSLSGSHTGESLPLATSNQVLIKFSAKGLAPARG 1759
CSMD3-201 FFQTSLHDVVEVYDGPTQQSSLLSSLSGSHSGESLPLSSGNQITIRFTSVGPITAKG 1862

CSMD1-217 FHFVYQAVPRTSDTQCSSVPEPRYGRRIGSEFSAGSIVRFECNPGYLLQGSTALHCQ 1782
CSMD2-204 FHFVYQAVPRTSATQCSSVPEPRYGKRLGSDFSVGAIVRFECNSGYALQGSPEIECL 1816
CSMD3-201 FHFVYQAVPRTSSTQCSSVPEPRFGRRIGNEFAVGSSVLFDCNPGYILHGSIAIRCE 1919

CSMD1-217 SVPNALAQWNDTIPSCVVPCSGNFTQRRGTILSPGYPEPYGNNLNCIWKIIVTEGSG 1839
CSMD2-204 PVPGALAQWNVSAPTCVVPCGGNLTERRGTILSPGFPEPYLNSLNCVWKIVVPEGAG 1873
CSMD3-201 TVPNSLAQWNDSLPTCIVPCGGILTKRKGTILSPGYPEPYDNNLNCVWKITVPEGAG 1976

CSMD1-217 IQIQVISFATEQNWDSLEIHDGGDVTAPRLGSFSGTTVPALLNSTSNQLYLHFQSDI 1896
CSMD2-204 IQIQVVSFVTEQNWDSLEVFDGADNTVTMLGSFSGTTVPALLNSTSNQLYLHFYSDI 1930
CSMD3-201 IQVQVVSFATEHNWDSLDFYDGGDNNAPRLGSYSGTTIPHLLNSTSNNLYLNFQSDI 2033

CSMD1-217 SVAAAGFHLEYKTVGLAACQEPALPSNSIKIGDRYMVNDVLSFQCEPGYTLQGRSHI 1953
CSMD2-204 SVSAAGFHLEYKTVGLSSCPEPAVPSNGVKTGERYLVNDVVSFQCEPGYALQGHAHI 1987
CSMD3-201 SVSAAGFHLEYTAIGLDSCPEPQTPSSGIKIGDRYMVGDVVSFQCDQGYSLQGHSHI 2090

CSMD1-217 SCMPGTVRRWNYPSPLCIATCGGTLSTLGGVILSPGFPGSYPNNLDCTWRISLPIGY 2010
CSMD2-204 SCMPGTVRRWNYPPPLCIAQCGGTVEEMEGVILSPGFPGNYPSNMDCSWKIALPVGF 2044
CSMD3-201 TCMPGPVRRWNYPIPICLAQCGGAMSDFSGVILSPGFPGNYPSSLDCTWTINLPIGF 2147

CSMD1-217 GAHIQFLNFSTEANHDFLEIQNGPYHTSPMIGQFSGTDLPAALLSTTHETLIHFYSD 2067
CSMD2-204 GAHIQFLNFSTEPNHDYIEIRNGPYETSRMMGRFSGSELPSSLLSTSHETTVYFHSD 2101
CSMD3-201 GVHLQFVNFSTETIHDYLEVRSGSSETSTVIGRLSGPQIPSSLFSTTHETSLYFHSD 2204

CSMD1-217 HSQNRQGFKLAYQAYELQNCPDPPPFQNGYMINSDYSVGQSVSFECYPGYILIGHPV 2124
CSMD2-204 HSQNRPGFKLEYQAYELQECPDPEPFANGIVRGAGYNVGQSVTFECLPGYQLTGHPV 2158
CSMD3-201 YSQNKQGFHIVYQAYQLQSCPDPRPFRNGFVIGNDFTVGQTISFECFPGYTLIGNSA 2261

CSMD1-217 LTCQHGINRNWNYPFPRCDAPCGYNVTSQNGTIYSPGFPDEYPILKDCIWLITVPPG 2181
CSMD2-204 LTCQHGTNRNWDHPLPKCEVPCGGNITSSNGTVYSPGFPSPYSSSQDCVWLITVPIG 2215
CSMD3-201 LTCLHGVSRNWNHPLPRCEALCGGNITAMNGTIYSPGYPDEYPNFQDCFWLVRVPPG 2318

CSMD1-217 HGVYINFTLLQTEAVNDYIAVWDGPDQNSPQLGVFSGNTALETAYSSTNQVLLKFHS 2238
CSMD2-204 HGVRLNLSLLQTEPSGDFITIWDGPQQTAPRLGVFTRSMAKKTVQSSSNQVLLKFHR 2272
CSMD3-201 NGIYINFTVLQTEPIYDFITVWDGPDQNSPQIGQFSGNTALESVYSTSNQILIKFHS 2375
```

CSMD1-217 DFSNGGFFVLNFHAFQLKKCQPPPAVPQAEMLTEDDDFEIGDFVKYQCHPGYTLVGT 2295
CSMD2-204 DAATGGIFAIAFSAYPLTKCPPPTILPNAEVVTENEEFNIGDIVRYRCLPGFTLVGN 2329
CSMD3-201 DFTTSGFFVLSYHAYQLRVCQPPPPVPNAEILTEDDEFEIGDIIRYQCLPGFTLVGN 2432

CSMD1-217 DILTCKLSSQLQFEGSLPTCEAQCPANEVRTGSSGVILSPGYPGNYFNSQTCSWSIK 2352
CSMD2-204 EILTCKLGTYLQFEGPPPICEVHCPTNELLTDSTGVILSQSYPGSYPQFQTCSWLVR 2386
CSMD3-201 AILTCRLGERLQMDGAPPVCQVLCPANELRLDSTGVILSPGYPDSYPNLQMCAWSIS 2489

CSMD1-217 VEPNYNITIFVDTFQSEKQFDALEVFDGSSGQSPLLVVLSGNHTEQSNFTSRSNQLY 2409
CSMD2-204 VEPDYNISLTVEYFLSEKQYDEFEIFDGPSGQSPLLKALSGNYSAPLIVTSSSNSVY 2443
CSMD3-201 VEKGYNITMFVEFFQTEKEFDVLQVYDGPNIQSPVLISLSGDYSSAFNITSNGHEVF 2546

CSMD1-217 LRWSTDHATSKKGFKIRYAAPYCSLTHPLKNGGILNRTAGAVGSKVHYFCKPGYRMV 2466
CSMD2-204 LRWSSDHAYNRKGFKIRYSAPYCSLPRAPLHGFILGQTSTQPGGSIHFGCNAGYRLV 2500
CSMD3-201 LQWSADHGNNKKGFRIRYIAFYCSTPESPPHGYIISQTGGQLNSVVRWACDRGFRLV 2603

CSMD1-217 GHSNATCRRNPLGMYQWDSLTPLCQAVSCGIPESPGNGSFTGNEFTLDSKVVYECHE 2523
CSMD2-204 GHSMAICTRHPQGYHLWSEAIPLCQALSCGLPEAPKNGMVFGKEYTVGTKAMYSCSE 2557
CSMD3-201 GKSSAVCRKSSYGYHAWDAPVPACQAISCGIPKAPTNGGILTTDYLVGTRVTYFCND 2660

CSMD1-217 GFKLESSQQATAVCQEDGLWSNKGKPPTCKPVACPSIEAQLSEHVIWRLVSGSLNEY 2580
CSMD2-204 GYHLQAGAEATAECLDTGLWSNRNVPPQCVPVTCPDVSSISVEHGRWRLIFETQYQF 2614
CSMD3-201 GYRLSSKELTTAVCQSDGTWSNHNKTPRCVVVTCPSINSFILEHGRWRIVNGSHYEY 2717

CSMD1-217 GAQVLLSCSPGYYLEGWRLLRCQANGTWNIGDERPSCRVISCGSLSFPPNGNKIGTL 2637
CSMD2-204 QAQLMLICDPGYYYTGQRVIRCQANGKWSLGDSTPTCRIISCGELPIPPNGHRIGTL 2671
CSMD3-201 KTKVVFSCDPGYHGLGPASIECLPNGTWSWRNERPYCQIISCGELPTPPNGNKIGTQ 2774

CSMD1-217 TVYGATAIFTCNTGYTLVGSHVRECLANGLWSGSETRCL------------------ 2676
CSMD2-204 SVYGATAIFSCNSGYTLVGSRVRECMANGLWSGSEVRCLATQTKLHSIFYKLLFDVL 2728
CSMD3-201 TSYGSTAIFTCDLGFMLVGSAVRECLSSGLWSESETRCL------------------ 2813

CSMD1-217 -------AGHCGSPDPIVNGHISGDGFSYRDTVVYQCNPGFRLVGTSVRICLQDHKW 2726
CSMD2-204 SSPSLTKAGHCGTPEPIVNGHINGENYSYRGSVVYQCNAGFRLIGMSVRICQQDHHW 2785
CSMD3-201 -------AGHCGIPELIVNGQVIGENYGYRDTVVYQCNPGFRLIGSSVRICQQDHNW 2863

CSMD1-217 SGQTPVCVPITCGHPGNPAHGFTNGSEFNLNDVVNFTCNTGYLLQGVSRAQCRSNGQ 2783
CSMD2-204 SGKTPFCVPITCGHPGNPVNGLTQGNQFNLNDVVKFVCNPGYMAEGAARSQCLASGQ 2842
CSMD3-201 SGQLPSCVPVSCGHPGSPIYGRTSGNGFNFNDVVTFSCNIGYLMQGPTKAQCQANRQ 2920

CSMD1-217 WSSPLPTCRVVNCSDPGFVENAIRHGQQNFPESFEYGMSILYHCKKGFYLLGSSALT 2840
CSMD2-204 WSDMLPTCRIINCTDPGHQENSVRQVHASGPHRFSFGTTVSYRCNHGFYLLGTPVLS 2899
CSMD3-201 WSHPPPMCKVVNCSDPGIPANSKRESKI-EHGNFTYGTVVFYDCNPGYFLFGSSVLI 2976

CSMD1-217 CMANGLWDRSLPKCLAISCGHPGVPANAVLTGELFTYGAVVHYSCRGSESLIGNDTR 2897
CSMD2-204 CQGDGTWDRPRPQCLLVSCGHPGSPPHSQMSGDSYTVGAVVRYSCIGKRTLVGNSTR 2956
CSMD3-201 CQPNGQWDKPLPECIMIDCGHPGVPPNAVLSGEKYTFGSTVHYSCTGKRSLLGQSSR 3033

CSMD1-217 VCQEDSHWSGALPHCTGNNPGFCGDPGTPAHGSRLGDDFKTKSLLRFSCEMGHQLRG 2954
CSMD2-204 MCGLDGHWTGSLPHCSGTSVGVCGDPGIPAHGIRLGDSFDPGTVMRFSCEAGHVLRG 3013
CSMD3-201 TCQLNGHWSGSQPHCSGDATGTCGDPGTPGHGSRQESNFRTKSTVRYACDTGYILHG 3090

CSMD1-217 SPERTCLLNGSWSGLQPVCEAVSCGNPGTPTNGMIVSSDGILFSSSVIYACWEGYKT 3011
CSMD2-204 SSERTCQANGSWSGSQPECGVISCGNPGTPSNARVVFSDGLVFSSSIVYECREGYYA 3070
CSMD3-201 SEERTCLANGSWTGRQPECKAVQCGNPGTTANGKVFRIDGTTFSSSVIYSCMEGYIL 3147

CSMD1-217 SGLMTRHCTANGTWTGTAPDCTIISCGDPGTLANGIQFGTDFTFNKTVSYQCNPGYV 3068
CSMD2-204 TGLLSRHCSVNGTWTGSDPECLVINCGDPGIPANGLRLGNDFRYNKTVTYQCVPGYM 3127
CSMD3-201 SGPSVRQCTANGTWSGTLPNCTIISCGDPGIPANGLRYGDDYVVGQNVSYMCQPGYT 3204

CSMD1-217 MEAVTSATIRCTKDGRWNPSKPVCKAVLCPQPPPVQNGTVEGSDFRWGSSISYSCMD 3125
CSMD2-204 MESHRVSVLSCTKDRTWNGTKPVCKALMCKPPPLIPNGKVVGSDFMWGSSVTYACLE 3184
CSMD3-201 MELNGSRIRTCTINGTWSGVMPTCRAVTCPTPPQISNGRLEGTNFDWGFSISYICSP 3261

CSMD1-217 GYQLSHSAILSCEGRGVWKGEIPQCLPVFCGDPGIPAEGRLSGKSFTYKSEVFFQCK 3182
CSMD2-204 GYQLSLPAVFTCEGNGSWTGELPQCFPVFCGDPGVPSRGRREDRGFSYRSSVSFSCH 3241
CSMD3-201 GYELSFPAVLTCVGNGTWSGEVPQCLPKFCGDPGIPAQGKREGKSFIYQSEVSFSCN 3318

CSMD1-217 SPFILVGSSRRVCQADGTWSGIQPTCIDPAHNTCPDPGTPHFGIQNSSRGYEVGSTV 3239
CSMD2-204 PPLVLVGSPRRFCQSDGTWSGTQPSCIDPTLTTCADPGVPQFGIQNNSQGYQVGSTV 3298
CSMD3-201 FPFILVGSSTRICQADGTWSGSSPHCIEPTQTSCENPGVPRHGSQNNTFGFQVGSVV 3375

CSMD1-217 FFRCRKGYHIQGSTTRTCLANLTWSGIQTECIPHACRQPETPAHADVRAIDLPTFGY 3296
CSMD2-204 LFRCQKGYLLQGSTTRTCLPNLTWSGTPPDCVPHHCRQPETPTHANVGALDLPSMGY 3355
CSMD3-201 QFHCKKGHLLQGSTTRTCLPDLTWSGIQPECIPHSCKQPETPAHANVVGMDLPSHGY 3432

CSMD1-217 TLVYTCHPGFFLAGGSEHRTCKADMKWTGKSPVCKSKGVR-E------VNETVTKTP 3346
CSMD2-204 TLIYSCQEGFSLKGGSEHRTCKADGSWTGKPPICLEVRPS-GRPINTAREPPLTQAL 3411
CSMD3-201 TLIYTCQPGFFLAGGTEHRVCRSDNTWTGKVPICEAGSKILVKDPRPALGTPSPKLS 3489

CSMD1-217 VPSDVFFVNSLWKGYYEYLGKRQPATLTVDWFNATSSKVNATFSEASPVELKLTGIY 3403
CSMD2-204 IPGDVFAKNSLWKGAYEYQGKKQPAMLRVTGFQVANSKVNATMIDHSGVELHLAGTY 3468
CSMD3-201 VPDDVFAQNYIWKGSYNFKGRKQPMTLTVTSFNASTGRVNATLSN-SNMELLLSGVY 3545

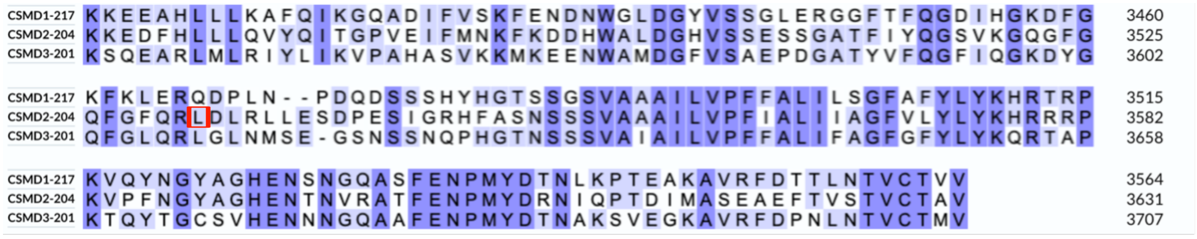


**Figure S1** Comparison of the CSMD1, CSMD2 and CSMD3 protein sequences. The residues conserved through two of the three paralogs are in light blue, while the ones conserved in all three of the paralogs are in dark blue. The biallelic mutated CSMD2 residues inherited in the affected individuals described in this report are highlighted in red, while the *de novo* mutations are highlighted in green. The residues reported mutated in references 31 and 32 and associated with epilepsy are in pink. The mutated CSMD1 residues identified in NDD individuals and reported in references 20 and 33 are highlighted in orange. This alignment was performed on the UniProt website1 using the MANE transcript of each protein (Homo_sapiens_ENSP00000489225_1 for CSMD1, Homo_sapiens_ENSP00000362479_4 for CSMD2 and Homo_sapiens_ENSP00000297405_5 for CSMD3).
