## Supplementary material for "Haploinsufficiency, *de novo* and biallelic missense variants in *CSMD2* are associated with neurodevelopmental disorders": Figure S2

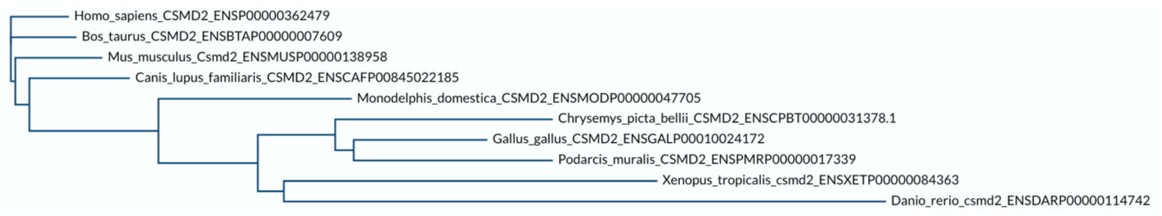


**Figure S2** Phylogenetic tree based on the alignment of the CSMD2 protein sequences. This alignment was performed on the UniProt website1.
