## Supplementary material for "Haploinsufficiency, *de novo* and biallelic missense variants in *CSMD2* are associated with neurodevelopmental disorders": Figure S3

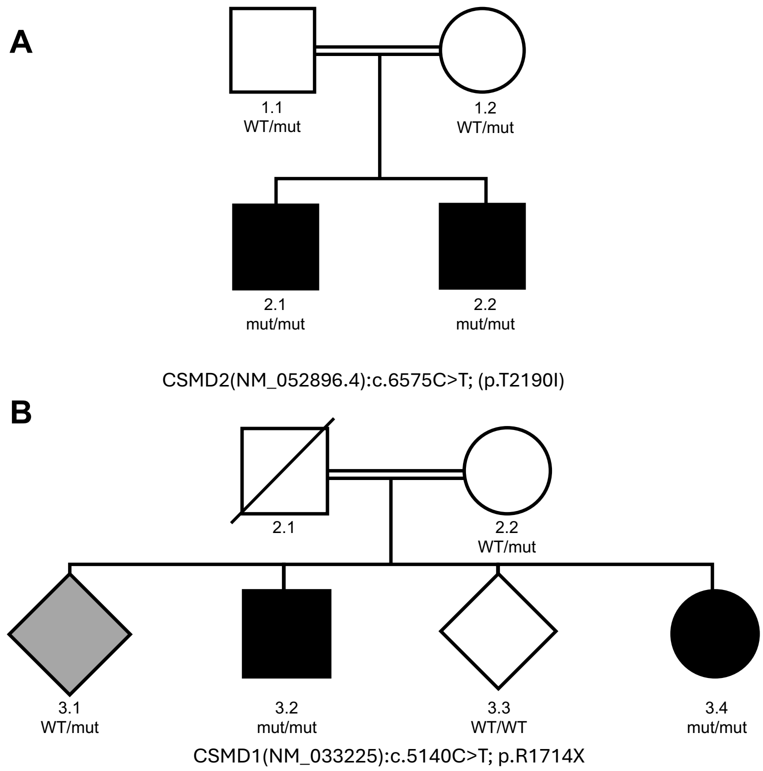


**Figure S3 Pedigrees. A.** Pedigree of the family segregating the CSMD2 p.(T2190I) missense variant. Phenotypes of the affected individuals are detailed in **Table S1**. **B.** Pedigree of the family segregating the CSMD1 p.(R1714X) stop-gain variant. Phenotypes of the affected individuals are detailed in **Table S6**.
