## Supplementary material for "Haploinsufficiency, *de novo* and biallelic missense variants in *CSMD2* are associated with neurodevelopmental disorders": Figure S4

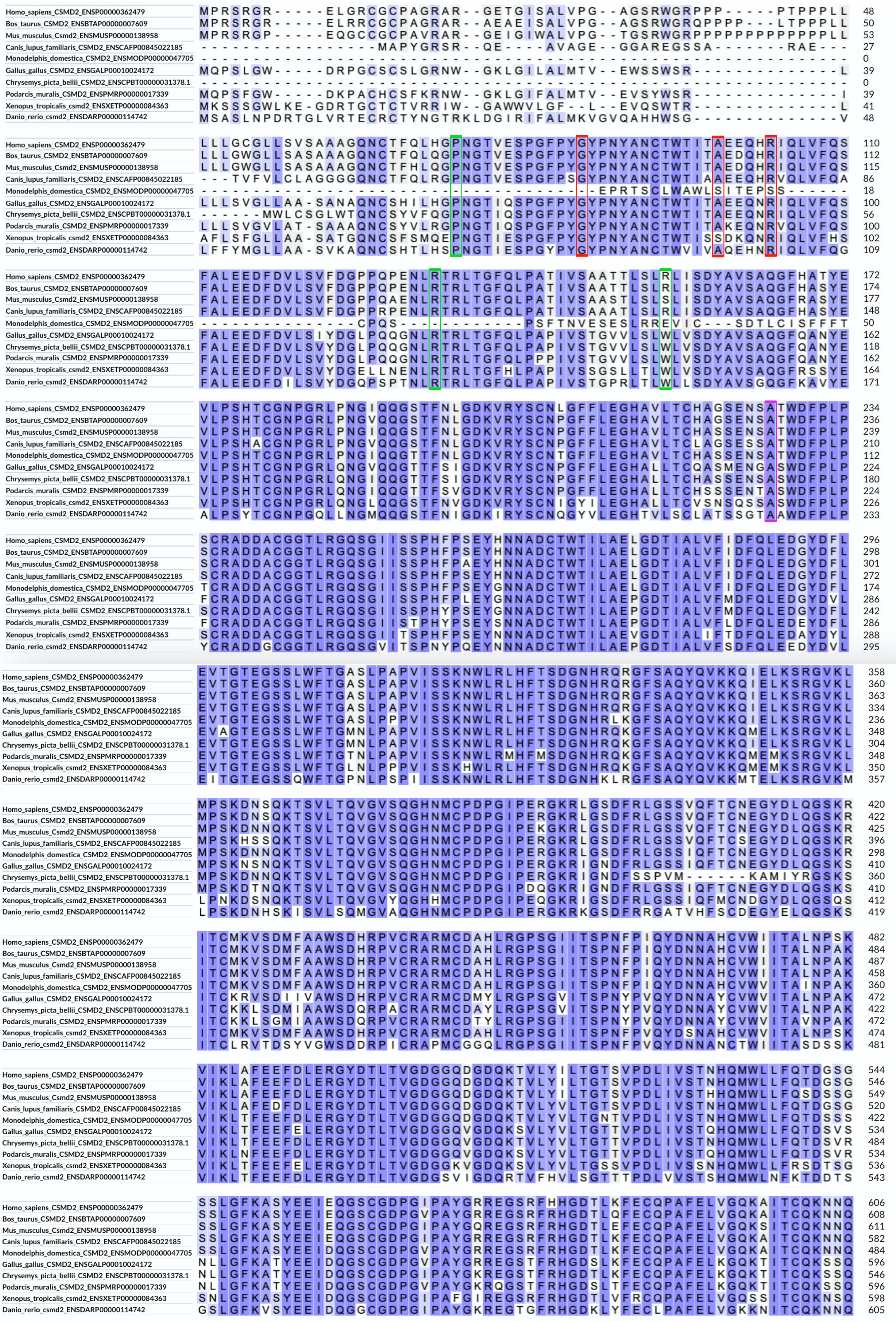

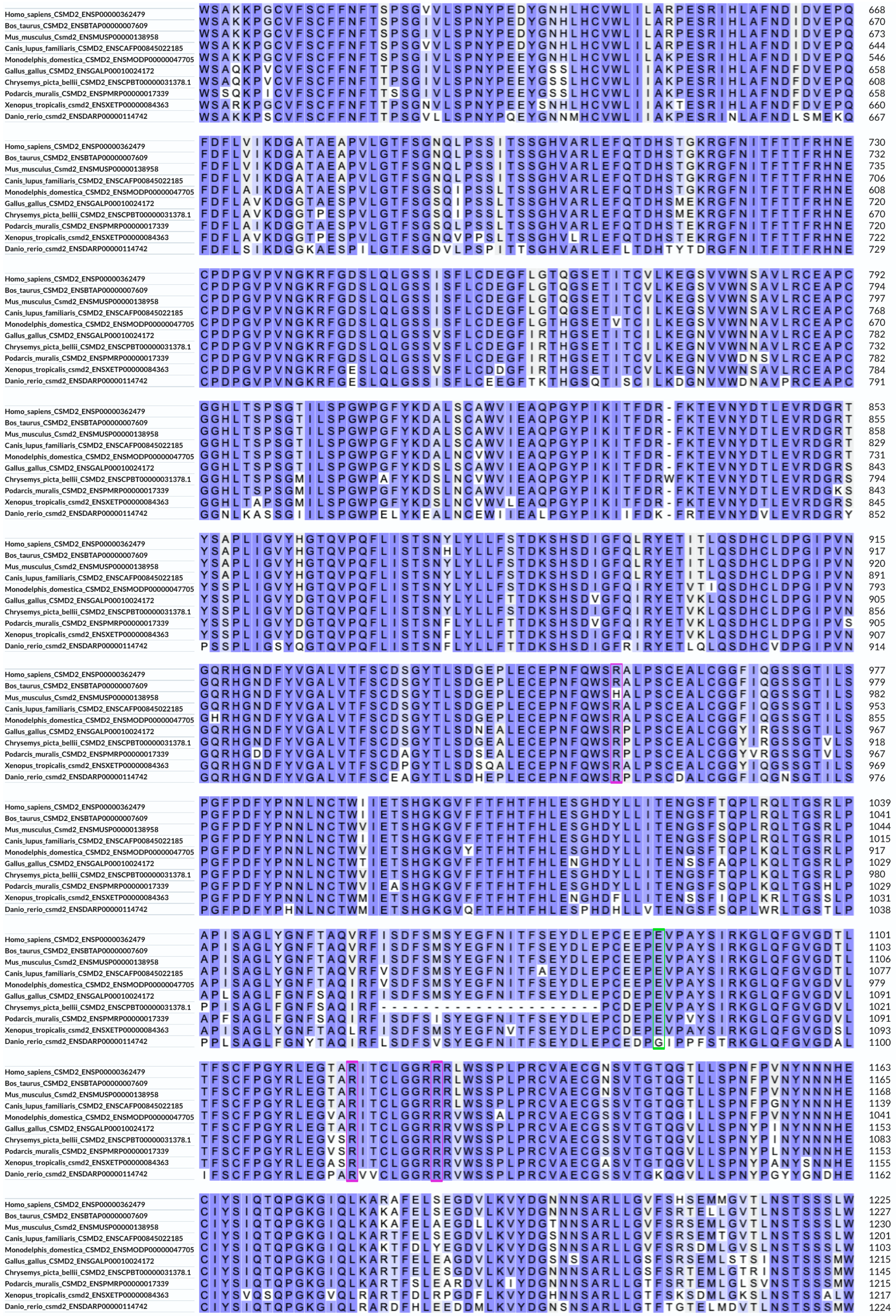
Homo_sapiens_CSMD2_ENSP00000362479
Bos_taurus_CSMD2_ENSBTAP00000007609
Mus_musculus_Csmd2_ENSMUSP00000138958
Canis_lupus_familiaris_CSMD2_ENSCAFP00845022185
Monodelphis_domestica_CSMD2_ENSMODP00000047705
Gallus_gallus_CSMD2_ENSGALP00010024172
Chrysemys_picta_bellii_CSMD2_ENSCPBT00000031378.1
Podarcis_muralis_CSMD2_ENSPMRP00000017339
Xenopus_tropicalis_csmd2_ENSXETP00000084363
Danio_rerio_csmd2_ENSDARP00000114742

```
Homo_sapiens_CSMD2_ENSP00000362479                LDFITDAENTSKGFELHFSSFELIKCEDPGTPKFGYKVHDEGHFAGSSVSFSCDPGYSLRGS 1287
Bos_taurus_CSMD2_ENSBTAP00000007609               LDFITDAENTSKGFELHFSSFELIKCEDPGTPQFGYKVHDGGHFAGSSVSFSCDPGYSLRGS 1289
Mus_musculus_Csmd2_ENSMUSP00000138958             LDFITDAENTSKGFELQFSSFELIRCEDPGTPQFGYKVHDGGHFAGSSVTFSCDPGYSLRGS 1292
Canis_lupus_familiaris_CSMD2_ENSCAFP00845022185   LDFITDADNTSKGFELHFSSFELIRCEDPGTPQFGYKVHDGGHFAGSSVSFSCDPGYSLRGS 1263
Monodelphis_domestica_CSMD2_ENSMODP00000047705    LDFITDSENTSKGFELQFSSFELIKCEDPGIPQFGYKVHDGGHFAGSSVSFSCDPGYTLRGS 1165
Gallus_gallus_CSMD2_ENSGALP00010024172            LEFITNSENTSKGFELQFSSFELIKCEDPGIPQFGYKVHDEGHFAGSSVSFNCDPGYTLRGS 1277
Chrysemys_picta_bellii_CSMD2_ENSCPBT00000031378.1 LEFISNSENTSKGFELQFSSFELIKCEDPGIPQFGYKVRDEGHFAGSSVSFNCDPGYTLRGS 1207
Podarcis_muralis_CSMD2_ENSPMRP00000017339         LEFISDGNSTSQGFELQFSSFELIKCEDPGIPQFGYKVHDEGHFAGSSVSFNCDPGYTLRGS 1277
Xenopus_tropicalis_csmd2_ENSXETP00000084363       LEFVSNSENTSKGFELYYSSFELIKCEDPGMPQYGYKEKDEGHFAGSSVTFSCDPGYSLRGS 1279
Danio_rerio_csmd2_ENSDARP00000114742              LEFISNSENTSKGFELHFTSFELVKCEDPGVPQFGFKREDKGHFAGSTVSYSCDPGYSLKGP 1286

Homo_sapiens_CSMD2_ENSP00000362479                EELLCLSGERRTWDRPLPTCVAECGGTVRGEVSGQVLSPGYPAPYEHNLNCIWTIEAEAGCT 1349
Bos_taurus_CSMD2_ENSBTAP00000007609               EELLCLSGERRAWDRPLPTCVAECGGTVRGEVSGQVLSPGYPAPYEHNLNCVWTIEADAGCT 1351
Mus_musculus_Csmd2_ENSMUSP00000138958             EELMCLSGERRTWDRPLPTCVAECGGTVRGEVSGQVLSPGYPAPYEHNLNCIWTIEADAGCT 1354
Canis_lupus_familiaris_CSMD2_ENSCAFP00845022185   EELLCLSGERRTWDRPLPTCVAECGGTVRGEVSGQVLSPGYPAPYEHNLNCIWTIEAEAGCT 1325
Monodelphis_domestica_CSMD2_ENSMODP00000047705    PVLMCLTGERRAWDRPLPICIAECGGTIKGEVSGQVLSPGYPAPYDHNLNCVWTIEADAGCT 1227
Gallus_gallus_CSMD2_ENSGALP00010024172            RVLVCLTGERRAWDQPLPTCVAECGGSIKGETSGRILSPGYPTPYDHNLNCIWSIEAEAGCT 1339
Chrysemys_picta_bellii_CSMD2_ENSCPBT00000031378.1 RALVCLTGERRAWDQPLPVCVAECGGTIRGESSGRILSPGYPTPYDHNLNCIWTIEAEAGCT 1269
Podarcis_muralis_CSMD2_ENSPMRP00000017339         RVLACLTGERRAWDQPLPICVAECGGSIKDEPSGRILSPGYPTPYDHNLNCVWTIEAEAGCT 1339
Xenopus_tropicalis_csmd2_ENSXETP00000084363       RVLMCLTGERRVWDQPLPVCVAECGGHIRGERVGRVLSPGYPAPYDHNLNCVWIIEADAGCT 1341
Danio_rerio_csmd2_ENSDARP00000114742              EVLTCLRGERRAWDSPLPQCVAECGGKIKDESMGRILSPGYPAPYEHNLHCVWTIEAALGST 1348

Homo_sapiens_CSMD2_ENSP00000362479                IGLHFLVFDTEEVHDVLRIWDGPVESGVLLKELSGPALPKDLHSTFNSVVLQFSTDFFTSKQ 1411
Bos_taurus_CSMD2_ENSBTAP00000007609               IGLHFLVFDTEEVHDVLRIWDGPVESGVLLKELSGPALPKDLHSTFNSVVLQFSTDFFTSKQ 1413
Mus_musculus_Csmd2_ENSMUSP00000138958             IGLHFLVFDTEEVHDVLRIWDGPVESGVLLKELSGPMLPKDLHSTFNSVVLQFSTDFFTSKQ 1416
Canis_lupus_familiaris_CSMD2_ENSCAFP00845022185   IGLHFLVFDTEEVHDVLRIWDGPVESGVLLKELSGAALPRDLHSTFSSVVLQFSTDFFTSKQ 1387
Monodelphis_domestica_CSMD2_ENSMODP00000047705    IGLHFLVFDTEEVHDVLRIWDGPVESGILLKELSGPMLPGNIHSTFSSVVLQFNTDFFTSKQ 1289
Gallus_gallus_CSMD2_ENSGALP00010024172            IGLHFMVFHTEEFHDVLRIWDGPVENGILLKEMSGSVLPSDVHSTFNSVILQFNTDFFTSKQ 1401
Chrysemys_picta_bellii_CSMD2_ENSCPBT00000031378.1 IGLHFMVFHTEEFHDVLRIWDGPE---------ISGSSLPSDVHSTFNSVVLQFNTDFFTSKQ 1323
Podarcis_muralis_CSMD2_ENSPMRP00000017339         IGLHFMVFHTEEFHDVLRIWDGPVEKGILLKEVSGSTLPSDVHSTFNSVVLQFNTDFFTSKQ 1401
Xenopus_tropicalis_csmd2_ENSXETP00000084363       IGLHLTVFHTEEVHDVLKIWDGSVESGILMKELSGSILPPDIHSTFNSVTLHFNTDFFTSKQ 1403
Danio_rerio_csmd2_ENSDARP00000114742              IGLHFLVFHTEEVHDVLRIWDGPQDGGVLLRELSGSTLPPDLHSTFNSVSLQFTTDFFTSKQ 1410

Homo_sapiens_CSMD2_ENSP00000362479                GFAIQFSVSTATSCNDPGIPQNGSRSGDSWEAGDSTVFQCDPGYALQGSAEISCVKIENRFF 1473
Bos_taurus_CSMD2_ENSBTAP00000007609               GFAIQFSVSTATSCNDPGVPQNGSRSGDSWEAGDSTVFQCDPGYALQGSAEISCVKIENRFF 1475
Mus_musculus_Csmd2_ENSMUSP00000138958             GFAIQFSVSTATSCNDPGVPQNGSRSGDSWEAGDSTVFQCDPGYALQGSAEISCVKIQNRFF 1478
Canis_lupus_familiaris_CSMD2_ENSCAFP00845022185   GFAIRFSVSTATSCNDPGVPQNGSRSGDSWEAGDSTVFQCDPGYALQGSAEISCVKIENRFF 1449
Monodelphis_domestica_CSMD2_ENSMODP00000047705    GFAIQFTVSTATSCNDPGVPQNGSRSGDSREAGDSIVFQCDPGYALQGMAQISCVKIENRFF 1351
Gallus_gallus_CSMD2_ENSGALP00010024172            GFTIQFSVSTATSCNDPGIPQNGSRSGDSKEAGDSIVFQCNPGYALQGEAKITCVQIENRFF 1463
Chrysemys_picta_bellii_CSMD2_ENSCPBT00000031378.1 GFAIQFSVSTATSCNDPGIPQNGSRSADSKEAGDSVVFQCDPGYTLQGEAKISCVQIENRFF 1385
Podarcis_muralis_CSMD2_ENSPMRP00000017339         GFAIQFSVSTATSCNDPGVPQNGSRSGDSKEAGDSIIFQCDPGYALQGDAKISCVQIENRFF 1463
Xenopus_tropicalis_csmd2_ENSXETP00000084363       GFSIHFSVTTATSCNDPGIPQNGSRSGDSRDAGDSIVFQCDPGYLLEGEPKISCVQIADRFY 1465
Danio_rerio_csmd2_ENSDARP00000114742              GFALQFS---ATSCNDPGIPTNGTRIGDSREPGDHVLFQCDPGYLLQGATKITCTEINNRFF 1469

Homo_sapiens_CSMD2_ENSP00000362479                WQPSPPTCIAPCGGDLTGPSGVILSPNYPEPYPPGKECDWKVTVSPDYVIALVFNIFNLEPG 1535
Bos_taurus_CSMD2_ENSBTAP00000007609               WQPSPPTCIAPCGGDLTGPSGVILSPNYPEPYPPGKECDWKVTVSPDYVIALVFNIFNLEPG 1537
Mus_musculus_Csmd2_ENSMUSP00000138958             WQPSPPTCIAPCGGDLTGPSGVILSPNYPEPYPPGKECDWKVTVSPDYVIALVFNTFNLEPG 1540
Canis_lupus_familiaris_CSMD2_ENSCAFP00845022185   WQPSPPTCIAPCGGDLTGPSGVILSPNYPEPYPPGKECDWRVTVSPDYVIALVFNTFNLEPG 1511
Monodelphis_domestica_CSMD2_ENSMODP00000047705    WQPDPPTCIAPCGGDLTGPSGVILSPNYPEPYPPGKECDWKVTVSPDYVIALVFNTFNLEPG 1413
Gallus_gallus_CSMD2_ENSGALP00010024172            WQPDPPSCTAPCGGILTGPAGVILSPQYPEPYPPGKECDWKLTVSPDYVIALVFNIFSLEPG 1525
Chrysemys_picta_bellii_CSMD2_ENSCPBT00000031378.1 WQPDPPSCIAPCGGILTGPAGVILSPQYPEPYPPGKECDWKLTVSPDYVIALVFNIFNLEPG 1447
Podarcis_muralis_CSMD2_ENSPMRP00000017339         WQPDPPTCIAPCGGILTGPSGVILSPNYPEPYPPGKECDWKLTVSPDYVIALVFHTFNLEPG 1525
Xenopus_tropicalis_csmd2_ENSXETP00000084363       WNPNPPICIAPCGGNLGGPTGVILSPQYPEPYPPLKECDWKLTVSSDYVIALSFNIFSIEPG 1527
Danio_rerio_csmd2_ENSDARP00000114742              WQPDPPTCSAPCGGNLTGPSGLILSPEYPEPYPHGRECDWTVTVTLDHIISLNFNHFSLEPS 1531

Homo_sapiens_CSMD2_ENSP00000362479                YDFLHIYDGRDSLSPLIGSFYGSQLPGRIESSSNSLFLAFRSDASVSNAGFVIDYTENPRES 1597
Bos_taurus_CSMD2_ENSBTAP00000007609               YDFLHIYDGRDSLSPLIGSFYGSQLPGRIESSSNSLFLAFRSDASVSNTGFVIDYTENPRES 1599
Mus_musculus_Csmd2_ENSMUSP00000138958             YDFLHIYDGRDSLSPLIGSFYGSQLPSRIESSSNSLFLAFRSDASVSNAGFVIDYTENPRES 1602
Canis_lupus_familiaris_CSMD2_ENSCAFP00845022185   YDFLHIYDGRDSLSPLIGSFYGSQLPGRIESSSNSLFLAFRSDASVSSAGFVIDYTENPRES 1573
Monodelphis_domestica_CSMD2_ENSMODP00000047705    YDFLHIYDGLDSHSPLIGSFYGSQLPERIESSSNSLFLAFRSDASVSNLGFVIDYTENPRES 1475
Gallus_gallus_CSMD2_ENSGALP00010024172            YDFLHIYDGPDSLSPLIGSFYGSQLPERIESSSNSLFLAFRSDASVSNSGFVIEYTENPRES 1587
Chrysemys_picta_bellii_CSMD2_ENSCPBT00000031378.1 YDFLHIYDGPSSLSPLIGSFYGSQLPDRIESSSNNLFLAFRSDASVSNAGFVIDYTENPRES 1509
Podarcis_muralis_CSMD2_ENSPMRP00000017339         YDFLHIYDGPSSLSPLIGSFYDTQLPERIESSSNNLFLAFRSDASVSNMGFVIYYTENPRES 1587
Xenopus_tropicalis_csmd2_ENSXETP00000084363       YDFLHIYDGPDSLSPLIGSFYGSQLPERIESSSNSLFIAFRSDASVSSGGFVIDYTENPRES 1589
Danio_rerio_csmd2_ENSDARP00000114742              YDFLHIYDGPDSLSPLLGSFYGTDVPDRIESSSNTLFLAFRSDASLSSNGFVLQYTENPRES 1593

Homo_sapiens_CSMD2_ENSP00000362479                CFDPGSIKNGTRVGSDLKLGSSVTYYCHGGYEVEGTSTLSCILGPDGKPVWNNPRPVCTAPC 1659
Bos_taurus_CSMD2_ENSBTAP00000007609               CFDPGSIKNGTRVGSDLKLGSSITYYCHGGYEVEGSSTLSCILGPDGKPVWNHPRPVCTAPC 1661
Mus_musculus_Csmd2_ENSMUSP00000138958             CFDPGSIKNGTRVGSDLKLGSSVTYYCHGGYEVEGASTLSCILGPDGKPMWNNPRPVCTAPC 1664
Canis_lupus_familiaris_CSMD2_ENSCAFP00845022185   CFDPGSIKNGTRVGSDLKLGSSVTYYCHGGYEVQGTSTLSCVLGPDGKPAWNKPRPVCTAPC 1635
Monodelphis_domestica_CSMD2_ENSMODP00000047705    CFDPGSIKNGTRSGTDLKLGSTITYYCNGGYMIEGASTLTCVLAADGKPVWNKPRPICTAPC 1537
Gallus_gallus_CSMD2_ENSGALP00010024172            CFDPGSIKNGTRVGTDLKLGSTITYYCDGGYDIEGVSTLTCIMGEDGKPAWNKPRPTCTAPC 1649
Chrysemys_picta_bellii_CSMD2_ENSCPBT00000031378.1 CFDPGSVKNGTRVGTDLKLGSTITYYCDGGYEIEGVSTLTCIMGGDGKPAWNKPRPTCTAPC 1571
Podarcis_muralis_CSMD2_ENSPMRP00000017339         CFDPGSIKNGTRAGSELKLGSTITYYCGSGYVIDGVSTLTCIMGGDGKPVWNKPRPTCTAPC 1649
Xenopus_tropicalis_csmd2_ENSXETP00000084363       CFDPGTIKNGTRVGNDLKLGSTITYYCESGYEIQGTSTLTCMMGDDGKPIWNHPSPSCTAPC 1651
Danio_rerio_csmd2_ENSDARP00000114742              CFEPGLVRNGTRVGTELKLGATVTYYCDSGYTLEGDATLTCIMGSDGKPGWNKPKPVCIAPC 1655

Homo_sapiens_CSMD2_ENSP00000362479                GGQYVGSDGVVLSPNYPQNYTSGQICLYFVTVPKDYVVFGQFAFFHTALNDVVEVHDGHSQH 1721
Bos_taurus_CSMD2_ENSBTAP00000007609               GGQYVGSDGVVLSPNYPQNYTSGQICLYFVTVPKDYVVFGQFAFFHTALNDVVEVHDGHNQH 1723
Mus_musculus_Csmd2_ENSMUSP00000138958             GGQYVGSDGVVLSPNYPQNYTSGQTCLYFVTVPKDYVVFGQFAFFHTALNDIVEVHDGYSQH 1726
Canis_lupus_familiaris_CSMD2_ENSCAFP00845022185   GGQYVGSDGVVLSPNYPQNYTSGQICLYFVTVPKDYVVFGQFAFFHTALNDVVEVHDGHSPH 1697
Monodelphis_domestica_CSMD2_ENSMODP00000047705    GGQYVGSDGVVLSPNYPQNYTRGQVCLYFIMVPKDYVVFGQFAFFHTALNDVVEVHDGHNQH 1599
Gallus_gallus_CSMD2_ENSGALP00010024172            GGQYTGSDGVVLSPNYPQNYTSGQVCLYFIVVPKDYVVFGQFAFFQTALNDIIEVHDGPNEH 1711
Chrysemys_picta_bellii_CSMD2_ENSCPBT00000031378.1 GGQYTGSDGVVLSPNYPRNYTSGQICLYFVVVPKDYGICRAVSGLDSQL------------- 1620
Podarcis_muralis_CSMD2_ENSPMRP00000017339         GGQYTGSDGVVLSPNYPQNYTSGQVCLYFIVVPKDYVVFGQFAFFQTALHDIIEVHDGPNQH 1711
Xenopus_tropicalis_csmd2_ENSXETP00000084363       GGRYVSQEGVILSPNYPRNYTTGQSCSYFIVVPKDYVVFGQFTFFQTALNDVVEVHDGPDDQ 1713
Danio_rerio_csmd2_ENSDARP00000114742              GGQYSGLEGVVLSPGYPGNYSSGRTCLYSVVVPRDYVVFSQFAFFQTALNDVVEVYDGPTQH 1717

Homo_sapiens_CSMD2_ENSP00000362479                SRLLSSLSGSHTGESLPLATSNQVLIKFSAKGLAPARGFHFVYQAVPRTSATQCSSVPEPRY 1783
Bos_taurus_CSMD2_ENSBTAP00000007609               SRLLSSLSGSHTGESLPLATSNQVLVKFSAKGQVPARGFHFVYQAVPRTSATQCSSVPEPRY 1785
Mus_musculus_Csmd2_ENSMUSP00000138958             SRLLSSLSGSHTGESLPLATSNQVLIKFSAKGQVSARGFHFVYQAVPRTSATQCSSVPEPRY 1788
Canis_lupus_familiaris_CSMD2_ENSCAFP00845022185   ARLLSSLSGSHTGESLPLATSNQVLIKFSAKGQAPARGFHFVYQAVPRTSATQCSSVPEPRY 1759
Monodelphis_domestica_CSMD2_ENSMODP00000047705    SRLLSSLSGSHTGESLPLATSNQVLIKFSAKGQATARGFHFVYQAVPRTSATQCSSVPEPRY 1661
Gallus_gallus_CSMD2_ENSGALP00010024172            SRLLSSLSGSHTGESLPLATTNQILIKFSSKGQSTAKGFHFVYQAVPRTSATQCSSVPEPRY 1773
Chrysemys_picta_bellii_CSMD2_ENSCPBT00000031378.1 ------------------------------------------YRT----------------- 1623
Podarcis_muralis_CSMD2_ENSPMRP00000017339         SRLLSSLSGSHTGESLPLATTNQILIKFSAKGQATAKGFHFVYQAVPRTSATQCSSVPEPRY 1773
Xenopus_tropicalis_csmd2_ENSXETP00000084363       SRLLSSLSGSHTGESLPLATSNQILIKFSAKGQATAKGFHFVYQAVPRTSATQCSSIPEPKY 1775
Danio_rerio_csmd2_ENSDARP00000114742              ARVLSSLSGAHTGESLPLATSNQILIRFTSKGQSSSRGFHLVYQAVPRTSATQCSSVPEPRH 1779

Homo_sapiens_CSMD2_ENSP00000362479                GKRLGSDFSVGAIVRFECNSGYALQGSPEIECLPVPGALAQWNVSAPTCVVPCGGNLTERRG 1845
Bos_taurus_CSMD2_ENSBTAP00000007609               GRRLGSDFSVGAIVRFECNSGYALQGSPEIECLPVPGALAQWNVSAPTCVVPCGGNLTERRG 1847
Mus_musculus_Csmd2_ENSMUSP00000138958             GKRLGSDFSVGAIIRFECNSGYALQGSPEIECLPVPGALAQWNVSAPTCVVPCGGNLTERRG 1850
Canis_lupus_familiaris_CSMD2_ENSCAFP00845022185   GKRLGSDFSVGAVVRFDCNSGYALQGSPEIECLPVPGALAQWNVSAPTCVVPCGGNLTERRG 1821
Monodelphis_domestica_CSMD2_ENSMODP00000047705    GRRIGSDFSVGAVVHFECNSGYSLQGSHHIECLTVPGALAQWNVSVPICVVPCGGNLTERKG 1723
Gallus_gallus_CSMD2_ENSGALP00010024172            GRRLGSDFAVGAVVRFECSAGYVLQGSRSIECLTMPGALAQWNVSVPTCVVPCGGNLTERKG 1835
Chrysemys_picta_bellii_CSMD2_ENSCPBT00000031378.1 ----------------FC---------REQSCIGTEG------------------------- 1635
Podarcis_muralis_CSMD2_ENSPMRP00000017339         GRRIGSDFSVGAVIRFECIAGYALQGSHSIECLTMPGALAQWNSSVPTCVVPCGGNLTERKG 1835
Xenopus_tropicalis_csmd2_ENSXETP00000084363       GRRLGNDFSVGAVVRFECFPGYALQGSRNIECQAVMGALAQWNASVPSCVVPCGGNLTERKG 1837
Danio_rerio_csmd2_ENSDARP00000114742              GRRTGNNFAVGAVVSFECNAGYVLEGPSAIECLTVPNALAQWNGSMPSCIVPCGGNLTQRIG 1841
```

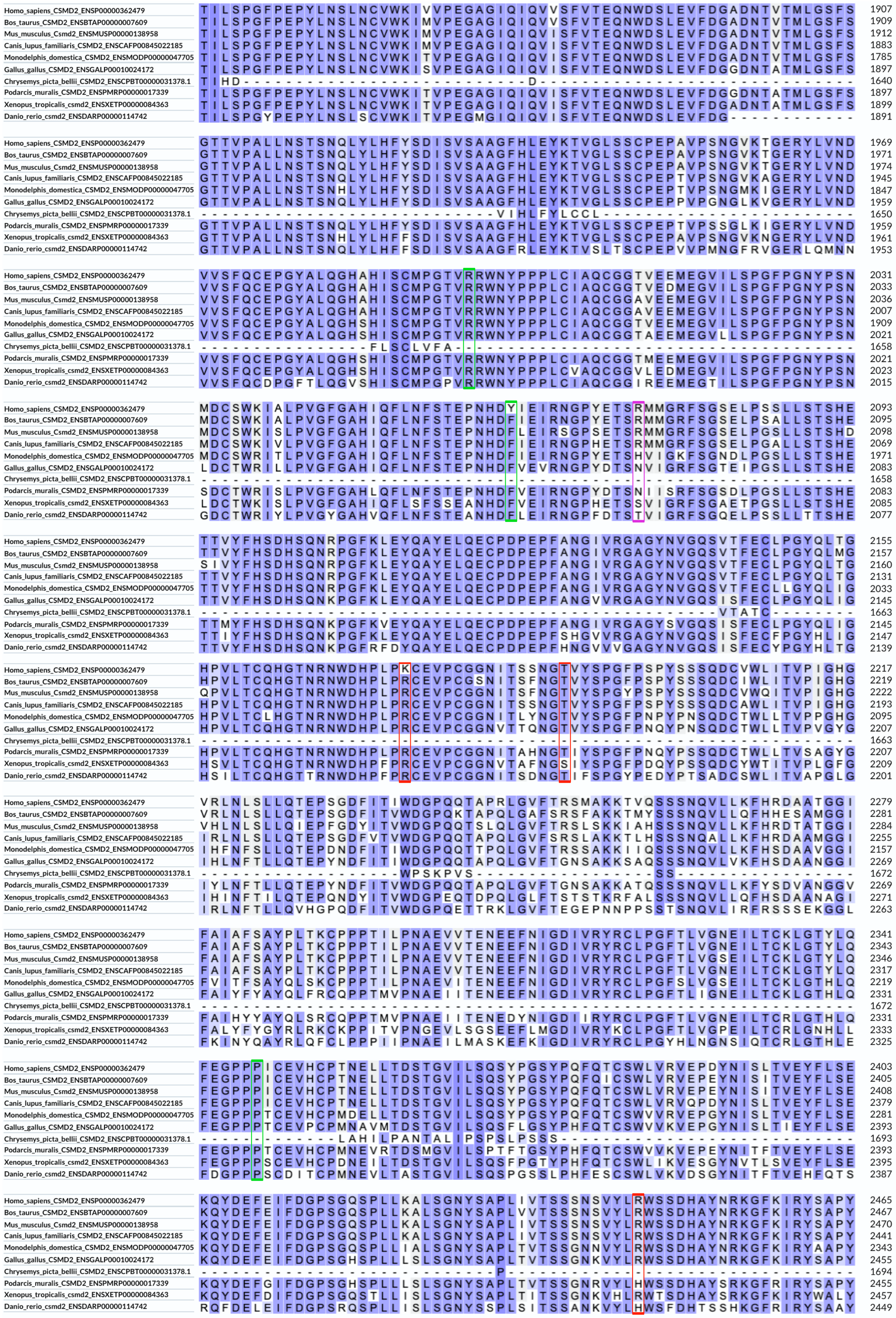

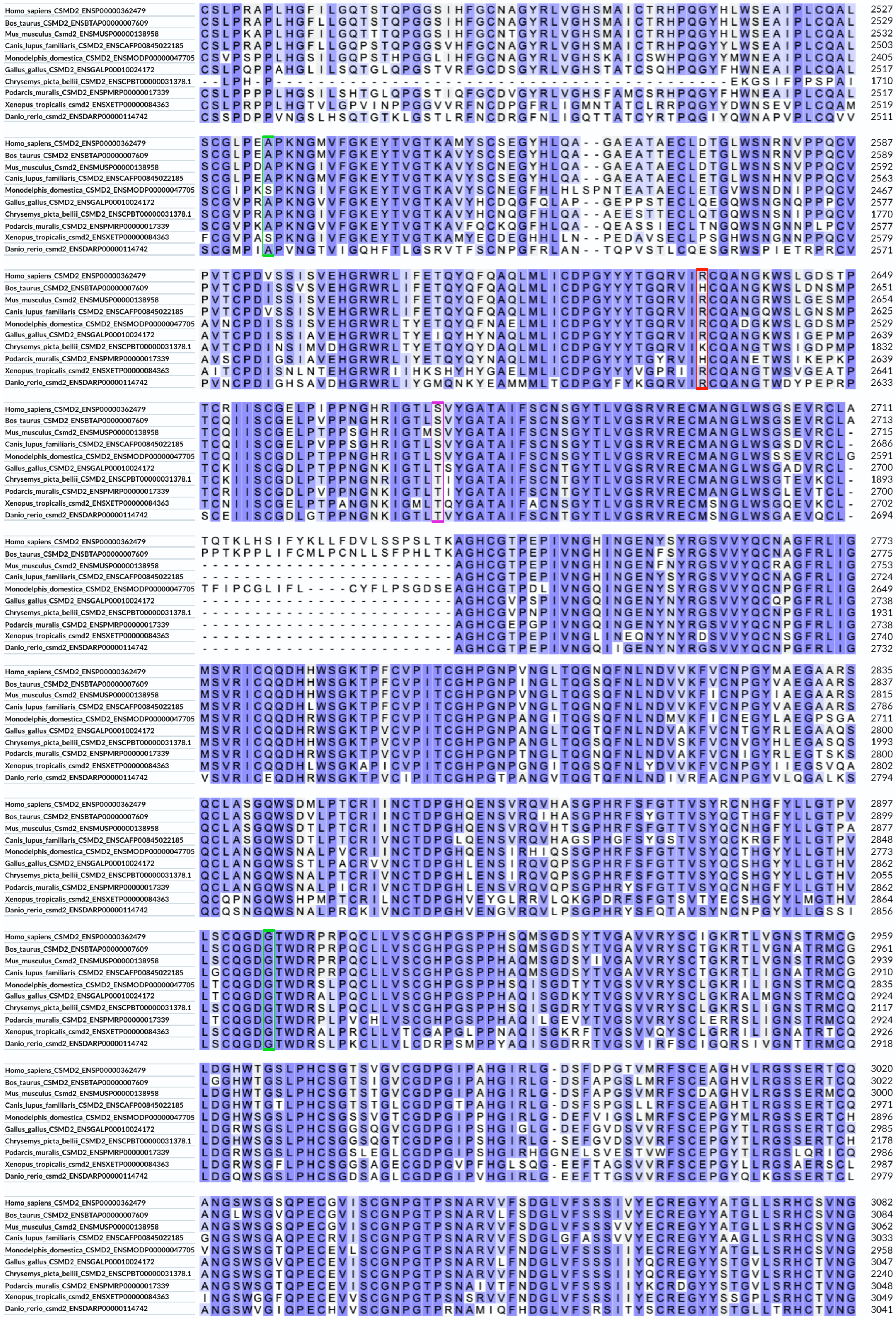

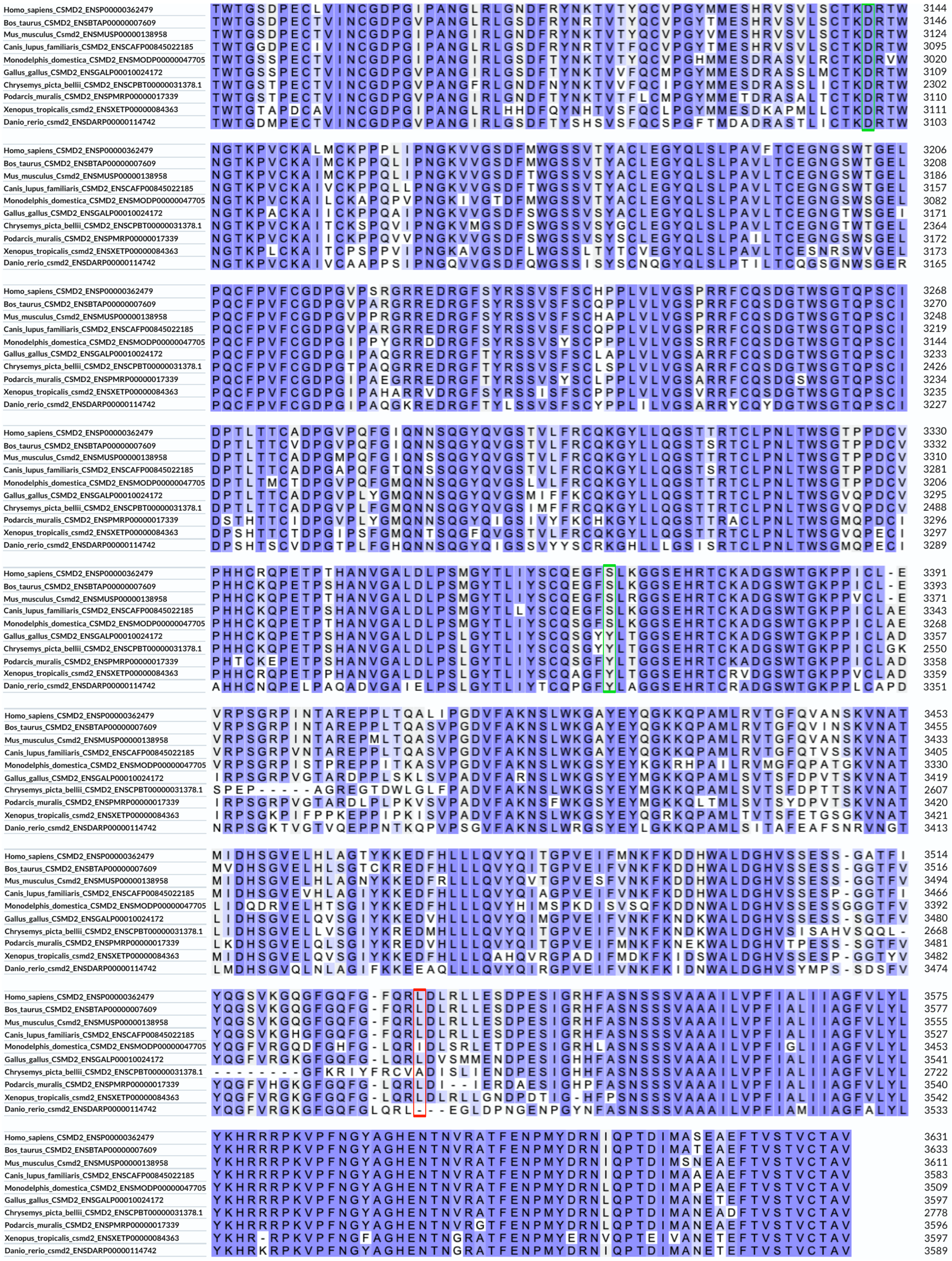

**Figure S4** Comparison of the CSMD2 protein sequences between ten vertebrate species. The darker the background of a residue, the more it is conserved. The biallelic mutated CSMD2 residues inherited in the affected individuals described in this report are highlighted in red, the *de novo* mutations are highlighted in green. The residues reported mutated in reference 31 and linked to epilepsy are in pink. This alignment was performed on the UniProt website1.
