## Supplementary material for "Haploinsufficiency, *de novo* and biallelic missense variants in *CSMD2* are associated with neurodevelopmental disorders": Figure S5

**A. CSMD1**

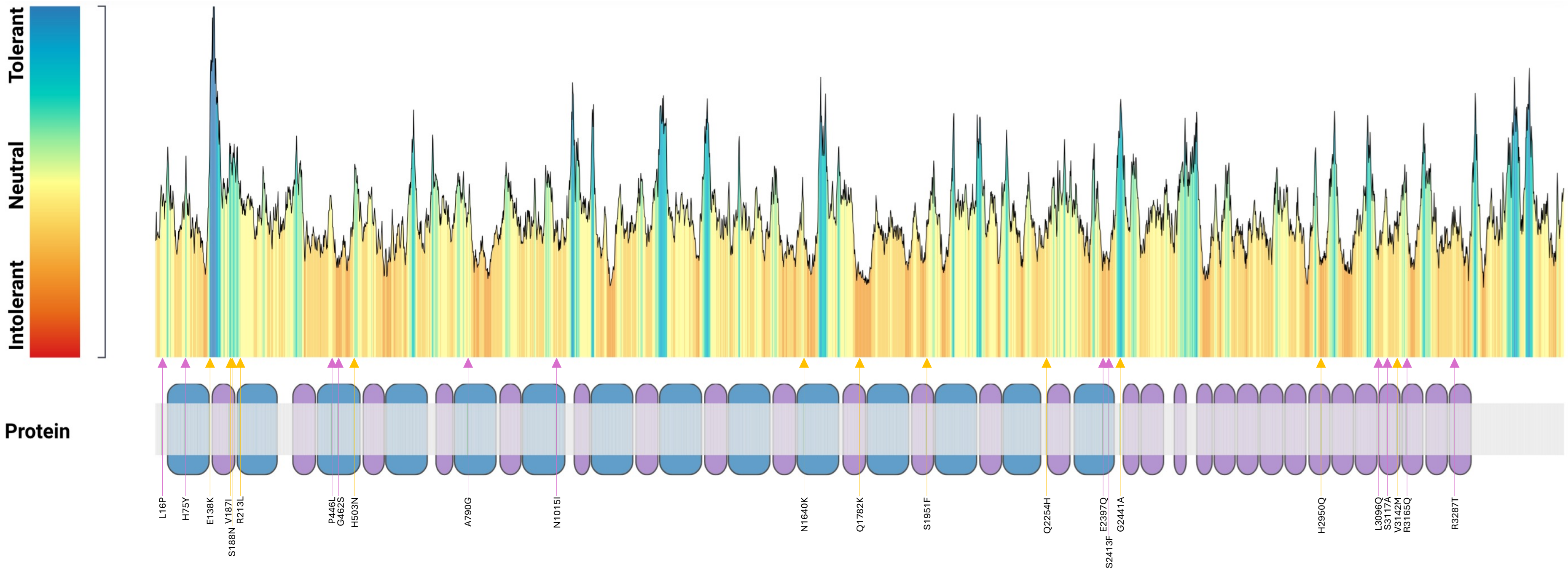
Tolerant
Neutral
Intolerant
Protein
L16P
H75Y
E138K
V187I
S188N
R213L
P446L
G462S
H503N
A790G
N1015I
N1640K
Q1782K
S1951F
Q2254H
E2397Q
S2413F
G2441A
H2950Q
L3096Q
S3117A
V3142M
R3165Q
R3287T

B. CSMD2

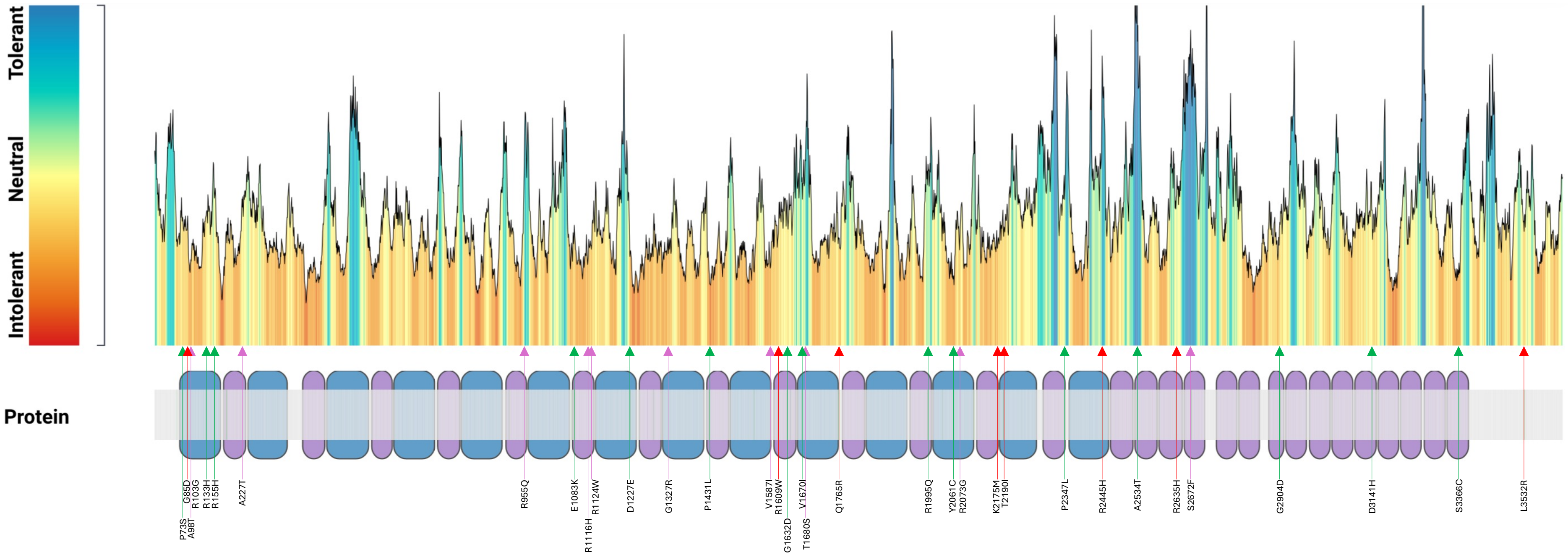
Tolerant
Neutral
Intolerant
Protein
P73S
G85D
A98T
R103G
R133H
R155H
A227T
R955Q
E1083K
R1116H
R1124W
D1227E
G1327R
P1431L
V1587I
R1609W
G1632D
V1670I
T1680S
Q1765R
R1995Q
Y2061C
R2073G
K2175M
T2190I
P2347L
R2445H
A2534T
R2635H
S2672F
G2904D
D3141H
S3366C
L3532R

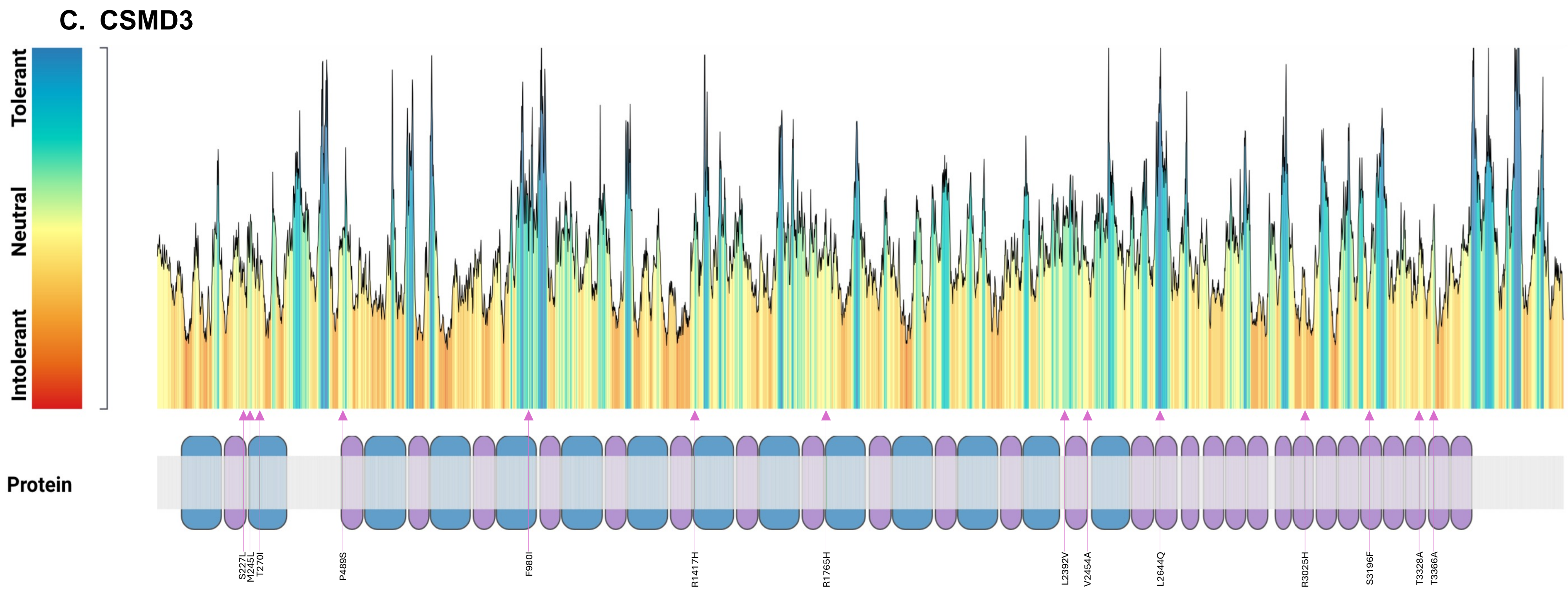


**Figure S5 Metadome diagrams.** Scheme showing the tolerance landscape for CSMD1 (**A**), CSMD2 (**B**) and CSMD3 (**C**) across species and across homologous human protein domains, respectively. The biallelic mutated CSMD2 residues inherited in the affected individuals described in this report are highlighted in red, the de novo mutations in green, residues reported mutated in references 31 and 32 and linked to epilepsy are in pink. The CSMD1 residues identified in NDD individuals and reported in references 20 and 33 are highlighted in orange.
