## Supplementary material for "Haploinsufficiency, *de novo* and biallelic missense variants in *CSMD2* are associated with neurodevelopmental disorders": Figure S6

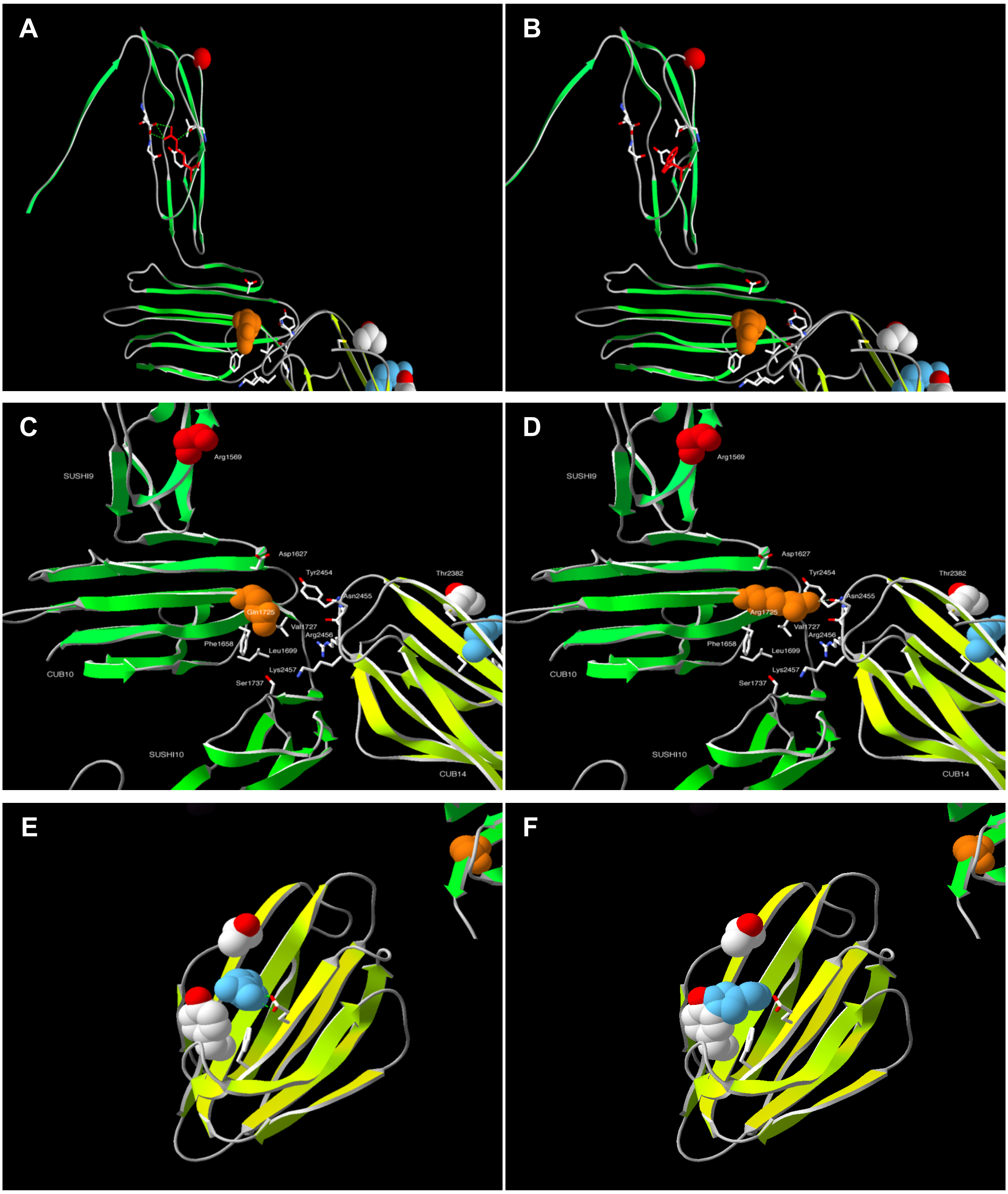


**Figure S6 Swiss-Pdb Viewer CSMD2 structure in ribbon representation. A.** Close-up view of R1609 of human CSMD2 protein. **B.** W1609 replacing the WT Arginine. **C.** Close -up view of Q1765 of human CSMD2 protein (Q1725 on AF-Q7Z408_v2 model). **D.** R1765 (R1725 on AF-Q7Z408_v2 model) replacing the WT Glutamine. **E.** Close-up view of R2445 of human CSMD2 protein. **F.** H2445 replacing the WT Arginine. Detailed descriptions are given in the text and **Table S3.**
