## Supplementary material for "Haploinsufficiency, *de novo* and biallelic missense variants in *CSMD2* are associated with neurodevelopmental disorders": Figure S7

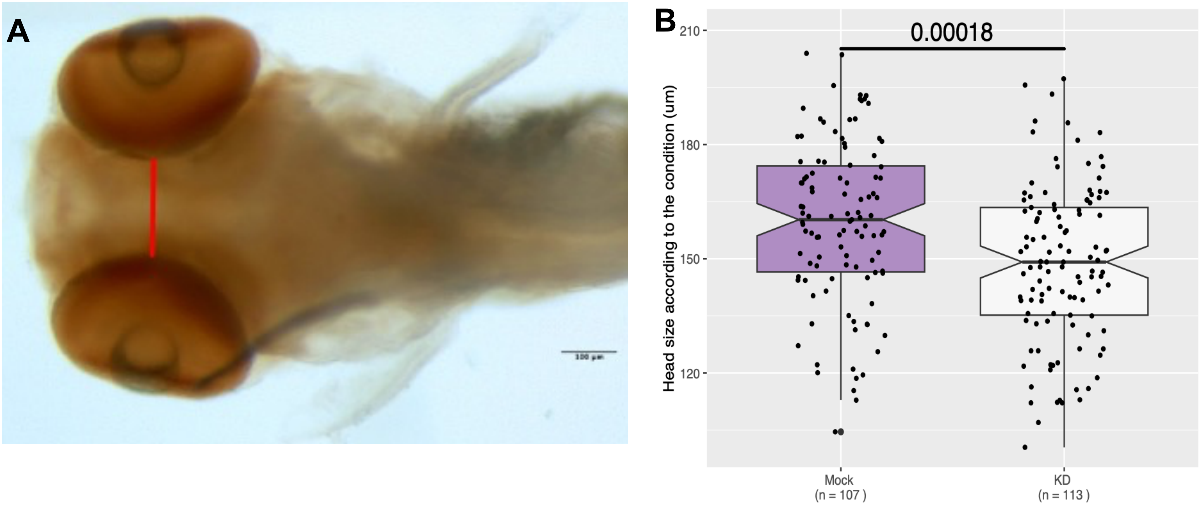


**Figure S7 Effects of *csmd2* knockdown on zebrafish head size A.** Picture of the head of a depigmented mock-injected larva. The red line shows where the head size was measured (magnification 6.3x). **B.** Head size at the level of the pupils in mock-injected and crispant larvae. Values below the 1st or above the 99th percentile were excluded as outliers prior to analysis. Boxes show the median and interquartile range, with notches indicating the 95% confidence interval around the median; whiskers represent the [Q1 - 1,5×IQR ; Q3 + 1,5×IQR] interval. Statistical significance between mock-injected and crispant groups was assessed using a two-sided Wilcoxon rank-sum test. Normalization was performed as detailed in Methods.
