## Supplementary material for "Haploinsufficiency, *de novo* and biallelic missense variants in *CSMD2* are associated with neurodevelopmental disorders": Figure S8

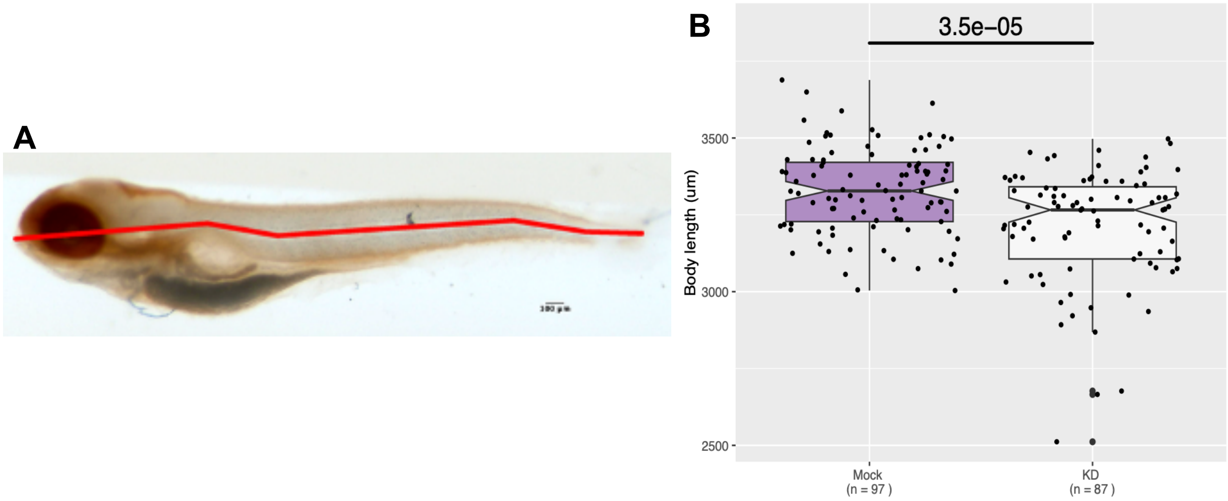


**Figure S8 Effects of *csmd2* knockdown on zebrafish body length A.** Picture of a depigmented mock-injected larva. The red segmented line shows body length measurement (magnification 2x). **B.** Body length, from the mouth to the tip of the tail in mock-injected and crispant larvae. Values below the 1st or above the 99th percentile were excluded as outliers prior to analysis. Boxes show the median and interquartile range, with notches indicating the 95% confidence interval around the median; whiskers represent the [Q1 - 1,5×IQR ; Q3 + 1,5×IQR] interval. Statistical significance between Mock and crispant groups was assessed using a two-sided Wilcoxon rank-sum test. Normalization was performed as detailed in Methods.
