## Supplementary material for "Haploinsufficiency, *de novo* and biallelic missense variants in *CSMD2* are associated with neurodevelopmental disorders": Table S1

| Number | 1 | 2 | 3 | 4 | 5 | 6 | 7 | 8 | 9 |
|---|---|---|---|---|---|---|---|---|---|
| Gender | M | M | M | F | M | F | M | F | M |
| Variant on MANE (NM_001281956.2) | c.6569C>T ; p.(T2190I) | | c.48255C>G ; p.(R1609W) | c.254G>A ; p.(G85D) | c. 10595T>G ; p.(L3532R) | c.307A>G ; p.(R103G) | c.7334G>A ; p.(R2445H) paternal / c.5294A>G ; p.(Q1765R) maternal | c.6524A>T ; p.(K2175M) / c.7903C>T ; p.(R2635C) | c.292G>A ; p.(A98T) / c.5039C>G ; p.(T1680S) |
| Genotype | Homozygous | | Homozygous | Homozygous | Homozygous | Homozygous | Compound heterozygous | Compound heterozygous | Compound heterozygous |
| Variant inheritance | Inherited from heterozygous parents | | Inherited from heterozygous parents | Inherited from heterozygous parents | | | Inherited from heterozygous parents | Inherited from heterozygous parents | Inherited from heterozygous parents |
| Date/Year of birth/Age | 16-20 yo | 11-15 yo | 6-10 yo | 6-10 yo | 6-10 yo | 6-10 yo | 26-30 yo | 0-5 yo | 0-5 yo |
| Birth remarks | No (Normal) | No (Normal) | No (normal) | IUGR | Not available | Normal antenatal scans, term birth via SVD, immediate cry at birth, birth weight 2.8 kg | Congenital hydrocephalus | Born prematurely due to preterm labor/PROM, forceps assisted VD; 5 weeks in NICU | NA |
| Height (cm, SD) | 159 cm (≈ 0.4th percentile) | 158 cm (>98th percentile) | 114 cm (25-50th percentile) | 105 cm (50th percentile) | Not available | 110 cm (< 5th percentile) | 175 cm (≈ 50th percentile) | 94.4cm (65th percentile) | NA |
| Weight (cm, SD) | 55 kg (≈ 9th percentile) | 33 kg (25-50th percentile) | 23.3 kg (75th percentile) | 17,4 kg (50th percentile) | Not available | 15.5 kg (< 5th percentile) | 53 kg (≈ 2nd percentile) | 13.8kg (55th percentile) | NA |
| OFC (cm) | 54 cm (≈ 25th percentile) | 52 cm (25-50th percentile) | 54 cm (>97 centile) Macrocephaly before 5 yo | 49 cm (10th percentile) | Not available | 48.5cm | 62 cm (>97th percentile) | 47.8cm (18th percentile) | NA |
| Feeding difficulties | No | No | No | No | Not available | No | No | Delayed swallowing with episodic penetration but no aspiration; history of thickened liquids | NA |
| Facial dysmorphisms | No | No | Broad nasal tip. Lower columella | No | Low Set Ears, Open Mouth, Gaze unfocused | No | No | Flattened face and occiput | NA |
| Congenital malformations (limb anomalies) | No | No | Left incomplete cleft lip, microform right-sided cleft lip and complete bilateral cleft palate | No | No | NA | No | No | NA |

| | | | | | | | | | |
|---|---|---|---|---|---|---|---|---|---|
| **Epilepsy/seizure** | Generalized bilateral epileptiform activity. | Generalized bilateral epileptiform activity. | No | No | No | Yes, myoclonic jerks, facial twitching , blank staring episodes, EEG : normal | Normal EEG | No (no EEG done) | Seizure frequency : sGTCS, 6–7 times/d ; Seizure free duration : 2.5 y ; Effective ASMs : Oxcarazepine, Lacosamide ; EEG : left frontal and midline sharp-slow waves; left frontal-originating CPS |
| **Intellectual disability** | Mild ID | Mild ID | Yes | Global developmental delay | No | Moderate ID | Mild ID | Yes | Mild ID |
| **Speech and language development** | Normal | Normal | Nonverbal | Speech delay | Speaking difficulty | Delayed | Normal | Vocalizes but no single words, uses some gestures to communicate | NA |
| **Learning and behavior** | Learning difficulties | Learning difficulties | Learning difficulties | Learning difficulties | History of loss of memory | Repetitive behaviors such as hand flapping, eye rolling, and tremors | Learning difficulties | Learning difficulties | NA |
| **Autism** | Yes (severe) | Yes (mild) | Yes(severe) | Yes | No | Yes (mild) | No | Suspected- no formal evaluation | NA |
| **ADHD** | Yes | Yes | No formal diagnosis | Yes | No | Yes | No | No | NA |
| **Vision and hearing** | Normal | Normal | Normal | Normal | Not available | Normal | Myopia, astigmatism, mild strabismus | B/l hyperopia with astigmatism (no correction); no hearing concerns | NA |
| **Motor development** | Normal | Normal | Normal | Normal | Normal | Delayed | Psychomotor delay | Delayed | NA |
| **Muscle tone (hypotonia, ...)** | Normal | Normal | Normal | Normal | Normal | Hypermobile joints | Normal | Generalized hypotonia | NA |
| **Ataxia/spasticity** | No | No | No | No | Not available | No | Mild motor incoordination | History of "wide based gait"; but not anymore | NA |

| | | | | | | | | | |
|---|---|---|---|---|---|---|---|---|---|
| **Brain anomalies** | Generalized brain atrophy with expansion of CSF convexities. | Generalized brain atrophy with expansion of CSF convexities. | No brain MRI | NA | Multiple axial slices were obtained through the brain from the base of the skull to the vertex. Bilateral symmetrical subtle increase in hypodensity in white matter. | Terminal zone of myelination in the periventricular region of the posterior horns of bilateral lateral ventricles (seen before 5yo, follow-up advised). Small midline epidermal inclusion cyst in the frontal bone | Rhomboence-phalosynapsis and severely dilated cerebral ventricles | 2 weeks: increased periventricular echogenicity at US. 4 weeks : head US normal; no brain MRI | Focal cortical dysplasia |
| **Skeletal anomalies** | No | No | Mild Pectus excavatum with chest asymmetry | No | No | Pes planus | No | No | NA |
| **Other anomalies** | Nothing | Nothing | Nothing | Nothing | Leukodystrophy (LDD) | Stage II CKD ( GFR 6.05 ml/min/1.73, BUN 15 mg/dl, Creatinine 1 mg/dl). Additional variant: NPHP4: NM_015102: c.1149_ 1150insCC: p.A384Pfs*23. | Inguinal hernia | Mild asthma, atopic dermatitis | NA |

**Table S1 Genotype and phenotype of individuals with *CSMD2* biallelic missense variants**
