## Supplementary material for "Haploinsufficiency, *de novo* and biallelic missense variants in *CSMD2* are associated with neurodevelopmental disorders": Table S2

| p. MANE (NM_001 281956.2) | State | Genomic variant position | c. MANE (NM_001 281 956.2) | Allele frequency (gnomAD total) | GERP ++ RS (CADD. gs) | PhyloP vertebrates (CADD.gs) | CADD PHRED (CADD. gs) | SIFT (CADD.gs) | PolyPhen (CADD.gs) | AlphaMissense (OpenCRAVAT) | MetaDome tolerance | Reference |
|---|---|---|---|---|---|---|---|---|---|---|---|---|
| P73S | *De novo* heterozygous missense | chr1:34089164 G>A | c.217C>T | NA | 6.54 | 11.295 | 24.9 | 0.07; tolerated | 0.985; probably_ damaging | 0.1406; likely_benign | Slightly intolerant (0.68) | Reported in this study |
| G85D | Homozygous biallelic missense | chr1:34089127 C>T | c.254G>A | NA | 6.54 | 7.518 | 27.6 | 0; deleterious | 1; probably_ damaging | 0.9794; likely_pathogenic | Intolerant (0.36) | Reported in this study |
| A98T | Compound heterozygous biallelic missense (3) | chr1:34089089 C>T | c.292G>A | 0.0001041 | 6.54 | 3.017 | 20.5 | 0.39; tolerated | 0.083; benign | 0.1025; likely_benign | Slightly intolerant (0.53) | Li *et al.*, 2025 |
| R103G | Homozygous biallelic missense | chr1:34089074 T>C | c.307A>G | 0.00001363 | 1.8 | 2.051 | 21.5 | 0.06; tolerated | 0.999; probably_ damaging | 0.9422; likely_pathogenic | Intolerant (0.52) | Reported in this study |
| R133H | *De novo* heterozygous missense | chr1:34088983 C>T | c.398G>A | 0.00004525 | 5.51 | 4.648 | 24.1 | 0; deleterious | 0.867; possibly_ damaging | 0.2541; likely_benign | Slightly intolerant (0.68) | Kaplanis *et al.*, 2020 |
| R155C | *De novo* heterozygous missense | chr1:34032648 G>A | c.463C>T | 0.00004743 | 5.59 | 4.518 | 25.1 | 0.02; deleterious | 0.003; benign | 0.5714; likely_pathogenic | Neutral (0.78) | Epi4K Consortium & Epilepsy Phenome / Genome Project, 2013 |
| E1083K | *De novo* heterozygous missense | chr1:33714746 C>T | c.3247G>A | 0.00001363 | 6.54 | 7.518 | 28.9 | 0.02; deleterious | 0.577; possibly_ damaging | 0.1278; likely_benign | Slightly intolerant (0.64) | Kaplanis *et al.*, 2020 |
| D1227E | *De novo* heterozygous missense | chr1:33700569 A>C | c.3681T>G | NA | -1.25 | -0.005 | 9.035 | 1; tolerated | 0.009; benign | 0.1485; likely_benign | Intolerant (0.49) | Kaplanis *et al.*, 2020 |
| P1431L | *De novo* heterozygous missense | chr1:33658101 G>A | c.4292C>T | 0.00003408 | 6.53 | 11.29 | 31 | 0; deleterious | 0.847; possibly_ damaging | 0.5998; likely_pathogenic | Intolerant (0.37) | Kaplanis *et al.*, 2020 |
| R1609W | Homozygous biallelic missense | chr1:33636504 G>A | c.4825C>T | 0.0000285 | 4.49 | 5.811 | 24.3 | 0; deleterious | 0.999; probably_ damaging | 0.4613; ambiguous | Slightly intolerant (0.67) | Reported in this study |
| G1632D | *De novo* heterozygous missense | chr1:33636434 C>T | c.4895G>A | NA | 6.22 | 5.693 | 21.5 | 0.11; tolerated | 0.105; benign | 0.1746; likely_benign | Neutral (0.74) | Reported in this study |
| V1670I | *De novo* heterozygous missense | chr1:33635292 C>T | c.5008G>A | 6.20E-07 | 6.54 | 5.84 | 25.1 | 0.03; deleterious | 0.924; probably_ damaging | 0.0798; likely_benign | Slightly tolerant (0.89) | Kaplanis *et al.*, 2020 |
| T1680S | Compound heterozygous | chr1:33635261 G>C | c.5039C>G | NA | 4.66 | 3.892 | 18.23 | 0.46; tolerated | 0.26; benign | 0.078; likely_benign | Tolerant (1.09) | Li *et al.*, 2025 |

| | | | | | | | | | | | | |
|---|---|---|---|---|---|---|---|---|---|---|---|---|
| | biallelic missense (3) | | | | | | | | | | | |
| Q1765R | Compound heterozygous biallelic missense (1) | chr1:33626488 T>C | c.5294A>G | NA | 6.54 | 8.871 | 32 | 0; deleterious | 0.999; probably_ damaging | 0.3997; ambiguous | Intolerant (0.29) | Reported in this study |
| R1995Q | *De novo* heterozygous missense | chr1:33616938 C>T | c.5984G>A | 0.000002478 | 6.54 | 7.518 | 30 | 0; deleterious | 0.999; probably_ damaging | 0.5859; likely_pathogenic | Slightly tolerant (0.96) | Khan *et al.*, 2024 |
| Y2061C | *De novo* heterozygous missense | chr1:33611202 T>C | c.6182A>G | 0.000003098 | 4.02 | 2.403 | 24.0 | 0; deleterious | 0.829; possibly_ damaging | 0.845; likely_pathogenic | Intolerant (0.37) | Kaplanis *et al.*, 2020 |
| K2175M | Compound heterozygous biallelic missense (2) | chr1:33605290 T>A | c.6524A>T | NA | 0.667 | 0.897 | 21.3 | 0; deleterious | 0.705; possibly_ damaging | 0.2215; likely_benign | Intolerant (0.31) | Reported in this study |
| T2190I | Homozygous biallelic missense | chr1:33602510 G>A | c.6569C>T | 0.000001245 | 6.54 | 11.295 | 26.6 | 0; deleterious | 0.979; probably_ damaging | 0.3539; ambiguous | Intolerant (0.36) | Reported in this study |
| P2347L | *De novo* heterozygous missense | chr1:33586515 G>A | c.7040C>T | 0.00001553 | 6.24 | 11.127 | 27.7 | 0; deleterious | 1; probably_ damaging | 0.9061; likely_pathogenic | Neutral (0.77) | Kaplanis *et al.*, 2020 |
| R2445H | Compound heterozygous biallelic missense (1) | chr1:33580806 C>T | c.7334G>A | 0.00006071 | 6.47 | 3.621 | 24.0 | 0.28; tolerated | 0.077; benign | 0.102; likely_benign | Tolerant (1.07) | Reported in this study |
| A2534T | *De novo* heterozygous missense | chr1:33572668 C>T | c.7600G>A | NA | 6.54 | 1.438 | 23.5 | 0.05; tolerated | 0.605; possibly_ damaging | 0.0862; likely_benign | Highly tolerant (2) | Kaplanis *et al.*, 2020 |
| R2635C | Compound heterozygous biallelic missense (2) | chr1:33571586 G>A | c.7903C>T | 0.00001032 | 6.54 | 4.71 | 33 | 0; deleterious | 0.972; probably_ damaging | 0.2233; likely_benign | Slightly intolerant (0.54) | Reported in this study |
| D3141H | *De novo* heterozygous missense | chr1:33541166 C>G | c.9421G>C | NA | 6.52 | 5.759 | 27.2 | 0.02; deleterious | 0.992; probably_ damaging | 0.8211; likely_pathogenic | Slightly intolerant (0.56) | Lelieveld *et al.*, 2016 |
| S3366C | *De novo* heterozygous missense | chr1:33533124 G>C | c.10097C>G | NA | 6.38 | 5.752 | 25.0 | 0; deleterious | 0.202; benign | 0.4148; ambiguous | Intolerant (0.39) | Kaplanis *et al.*, 2020 |
| L3532R | Homozygous biallelic missense | chr1:33521465 A>C | c.10595T>G | 0.000006163 | 6.54 | 5.339 | 27.3 | 0.02; deleterious | 0.573; possibly_ damaging | 0.3188; likely_benign | Neutral (0.72) | Reported in this study |

**Table S2 Predicted deleteriousness of *CSMD2* variants**
