## Supplementary material for "Haploinsufficiency, *de novo* and biallelic missense variants in *CSMD2* are associated with neurodevelopmental disorders": Table S3

| p. MANE (NM_001281 956.2) | State | Chromosomal position | Domain 3D | 3D Prediction |
|---|---|---|---|---|
| P73S | *De novo* heterozygous | 1-34089164-G-A | CUB1 | Likely benign |
| G85D | Homozygous | 1-34089127-C-T | CUB1 | Introduction of a negative charge potentially affecting molecular interactions, effect on the domain's conformation, likely deleterious |
| A98T | Compound heterozygous (3) | 1-34089089-C-T | CUB1 | Clashes with surrounding residues, modification of the local packaging, likely deleterious |
| R103G | Homozygous | 1-34089074-T-C | CUB1 | Loss of a positive charge will likely disrupt the overall folding of the protein, likely deleterious |
| R133H | *De novo* heterozygous | 1-34088983-C-T | CUB1 | Disruption of long-range interaction, expected to be deleterious |
| R155C | *De novo* heterozygous | 1-34032648-G-A | CUB1 | Disruption of the molecular surface and charge of the domain, clashes, expected to be deleterious |
| E1083K | *De novo* heterozygous | 1-33714746-C-T | Sushi6 | Change of local negative charge to positive in a loop at the start of Sushi6, potentially disrupting affecting folding or protein-protein interactions |
| D1227E | *De novo* heterozygous | 1-33700569-A-C | CUB7 | Likely benign |
| P1431L | *De novo* heterozygous | 1-33658101-G-A | Sushi8 | Heavily clashes with surrounding residues, expected to disrupt the folding and be deleterious |
| R1609W | Homozygous | 1-33636504-G-A | Sushi9 | Loss of salt bridge and hydrogen bond, disruption of the fold |
| G1632D | *De novo* heterozygous | 1-33636434-C-T | CUB10 | Effect on the domain conformation, introduction of a negative charge potentially affecting molecular interactions, likely deleterious |
| V1670I | *De novo* heterozygous | 1-33635292-C-T | CUB10 | Heavily clashes with surrounding residues, expected to disrupt the folding and be deleterious |
| T1680S | Compound heterozygous (3) | 1-33635261-G-C | CUB10 | Likely benign |
| Q1765R | Compound heterozygous (1) | 1-33626488-T-C | CUB10 | Introduction of a positive charge which could perturb the interaction with CUB14 |
| R1995Q | *De novo* heterozygous | 1-33616938-C-T | Sushi11 | Probably tolerated |
| Y2061C | *De novo* heterozygous | 1-33611202-T-C | CUB12 | Structurally tolerated but an exposed Cys could lead to possible undesirable interactions and reactions with other molecules |
| K2175M | Compound heterozygous (2) | 1-33605290-T-A | Sushi12 | Disruption of salt-bridge and alteration of the surface's charge which could affect molecular interactions |
| T2190I | Homozygous | 1-33602510-G-A | CUB13 | Alteration of the protein surface (large hydrophobic residue instead of small polar one) and abolition of a possible phosphorylation site (NetPhos-3.1 prediction score of 0.827) in a strictly conserved residue context (NGTVYSPG, except in Xenopus: NGSVYSPG), potentially a major impact affecting the regulation or protein-protein interactions |
| P2347L | *De novo* heterozygous | 1-33586515-G-A | Sushi13 | In a 3 residues rigid poly-proline stretch, disruption of local folding, heavily clashes with surrounding residues, predicted to be very deleterious |
| R2445H | Compound heterozygous (1) | 1-33580806-C-T | CUB14 | Abolition of a salt bridge, clashes with surroundings residues, likely deleterious |
| A2534T | *De novo* heterozygous | 1-33572668-C-T | Sushi15 | Likely benign |
| R2635C | Compound heterozygous (2) | 1-33571586-G-A | Sushi16 | An exposed Cys could lead to possible undesirable interactions and reactions with other molecules |
| D3141H | *De novo* heterozygous | 1-33541166-C-G | Sushi22 | Introduction of a positive charge instead of a negative one, potential disruption of interactions with other domains of inside the protein, probably disruptive |
| S3366C | *De novo* heterozygous | 1-33533124-G-C | Sushi22 | Probably tolerated, with potential undesirable interactions and reactions with other molecules |
| L3532R | Homozygous | 1-33521465-A-C | Between last Sushi and transmembrane | Addition of a positive charge that might affect the structure and the stability |

**Table S3 3D model: position and predictive effects of the mutations observed in CSMD2**
