## Supplementary material for "Haploinsufficiency, *de novo* and biallelic missense variants in *CSMD2* are associated with neurodevelopmental disorders": Table S4

| Number | D1.2 | D1.3 | D2.2 | D3.1 | D3.2 | E2.1 | E2.2 | E2.3 |
|---|---|---|---|---|---|---|---|---|
| Gender | F | F | F | F | M | M | F | F |
| Variant on MANE (NM_001281956.2) | c.10328dup ; p.(Gln3444Profs*49) | NA | c.10328dup ; p.(Gln3444Profs*49) | c.10328dup ; p.(Gln3444Profs*49) | c.10328dup ; p.(Gln3444Profs*49) | c.4742_4743del ; p.(Val1581Glufs*8) | | |
| Genotype | Heterozygous | NA | Heterozygous | Heterozygous | Heterozygous | Homozygous | | |
| Date/Year of birth/Age | Adult | Adult | Adult | 11-15 yo | 11-15 yo | 15-20 yo | 15-20 yo | 6-10 yo |
| Birth remarks | None | None | None | None | None | Normal | Normal | Normal |
| Height (cm, SD) | NA | NA | NA | 150.8cm( -1.25SD) | 133cm ( -2.23SD) | 178 cm (-0.6 SD) | 161 cm (-1.2SD) | 121 cm (+0.6 SD) |
| Weight (cm, SD) | NA | NA | NA | 31.3kg ( -2.73SD) | 26.40kg (-2.23SD) | 76.2 kg (BMI 24.1 kg/m2 = +1.4SD) | 47.1 kg (BMI 18.1 = -1.1 SD) | 22.6 kg (BMI 15.4 kg/m2 = +0.2 SD) |
| OFC (cm, SD) | NA | NA | NA | NA | NA | 58 cm (+0.5 SD) | NA | Not measured |
| Feeding difficulties | No | No | No | No | No | No | No | No |
| Facial dysmorphisms | No | No | No | No | No | Mild synophris, broad nasal tip | NA | No |
| Congenital malformations (limb anomalies) | No | No | No | No | No | No | No | No |
| Epilepsy/seizure | No | Yes | No | No | No | No | No | No |
| Intellectual disability | No | Profound | No | No | Suspected | Mild ID, IQ 51 | NA | Mild ID, IQ 59 |
| Speech and language development | No | Speech regression around before 5 yo | No | Delayed ; First words on time but had delays in expressive language | Delayed ; First words on time, speech regression before 5 yo, slowly expanded vocabulary ; currently has receptive-expressive language disorder diagnosis | Delayed | NA | Delayed. Language production and comprehension below -3 SD, and development on the SON-R corresponding to a delay of 1 year and 3 months |
| Learning and behavior | No | No | Yes, learning difficulties | Yes, learning difficulties; receives some tutoring | Yes, learning difficulties; receives special education support | Learning difficulties | Learning difficulties | Learning difficulties |

| | | | | | | | | |
|---|---|---|---|---|---|---|---|---|
| **Autism** | Suspected | Yes | Yes | Yes | Yes; level 3 diagnosis | No | NA | No |
| **ADHD** | Suspected | No | Yes | Yes | Yes | No | NA | No |
| **Vision and hearing** | No | No | No | No | No | Normal | Normal | Normal |
| **Motor development** | No | NA | No | No | No | Delayed. Current motor problems but likely related to second diagnosis genetic sensorimotor polyneuropathy. *SORD* c.757del p.(Ala253Glnfs*27) | NA | Normal |
| **Muscle tone (hypotonia, ...)** | No | NA | No | No | Hypotonia | Dystonia and pain feet and legs (related to second diagnosis) | Normal | Normal |
| **Ataxia/spasticity** | No | NA | No | No | No | Has second diagnosis of genetic sensorimotor polyneuropathy | No | No |
| **Brain anomalies** | NA | NA | NA | NA | NA | Arachnoid cyst in the left retrocerebellar region. | NA | Unknown |
| **Skeletal anomalies** | No | No | No | No | No | No | No | No |
| **Other** | Anxiety, depression, OCD, hypermobility suspected | No | Hypermobility (hEDS diagnosis), OCD | Preauricular pit, hypermobility, OCD | OCD, hypermobility | Has second diagnosis of genetic sensorimotor polyneuropathy. Low threshold for nausea, e.g., when exposed to warm showers | Not seen in the clinic yet, has learning problems (has repeated 3 school years) | History of viral meningitis, liquor enterovirus + |

**Table S4 Genotype and phenotype of individuals with CSMD2 truncating variants**
