## Supplementary material for "Haploinsufficiency, *de novo* and biallelic missense variants in *CSMD2* are associated with neurodevelopmental disorders": Table S5

| Gender | F | F | F | M |
|---|---|---|---|---|
| **Variant on MANE (NM_001281956.2)** | c.4895G>A ; p.(Gly1632Asp) | c.217C>T ; p.(Pro73Ser) | c.6182A>G ; p.Tyr2061Cys | c.3681T>G ; p. Asp1227Glu |
| **Genotype** | Heterozygous | Heterozygous | Heterozygous | Heterozygous |
| **Variant inheritance** | *De novo* (paternity confirmed; Trio-Exome) | *De novo* | *De novo* | *De novo* |
| **Date/Year of birth/Age** | 11-25 yo | 6-10 yo | NA | 6-10 yo |
| **Birth remarks** | None | None | NA | Preeclampsia, child delivered by CS and not incubated |
| **Height (cm, SD)** | 149 cm (58th percentile) | 98 cm(+3SD) | NA | 71 cm (+03SD) (50-75thcentile) |
| **Weight (cm, SD)** | 40,8 kg (63th percentile) | 15,8 kg (+2,5SD) | NA | 5.2 kg (-4.3SD) (<1st centile) |
| **OFC (cm, SD)** | 53 cm (48th percentile) | 49,5 (+1SD) | NA | 42 cm (-2.14SD) (2nd centile) |
| **Feeding difficulties** | feeding difficulties in the first year of life, still picky eater | No | NA | Yes |
| **Facial dysmorphisms** | No | No | NA | No |
| **Congenital malformations (limb anomalies)** | No | No | NA | No |
| **Epilepsy/seizure** | No seizures, EEG 2017 normal | Theta and slow delta activity, spikes with rare myoclonies | Seizure | Seizures started before 5 yo as generalized tonic clonic without altered conscious level, last about 30 seconds and reached 4-5 times per day. EEG showed focal frontal epileptiform focus |
| **Intellectual disability** | Mild ID/ LD (Age 6-10 IQ: 71) | Mild ID | NA | Yes, couldn't respond to his name or recognize the mother, couldn't follow objects |
| **Speech and language development** | Delayed and impaired speech development; in early childhood only limited expressive language. Between age 11-15, the patient speaks full sentences, articulation is poor and sometimes indistinct | No language | NA | Not attained |
| **Learning and behavior** | Learning difficulties | Flapping | NA | NA |
| **Autism** | Yes (severe) | Yes | NA | Yes, recurrent turning head from onside to other, no eye contact |
| **ADHD** | No | Yes | NA | Excessive movement, itritable |
| **Vision and hearing** | Normal | No | NA | Couldn't follow by eyes or respond to his name but respond to voices, |

| | | | | |
|---|---|---|---|---|
| | | | | normal fundus examination and ABR |
| **Motor development** | Normal | No | NA | Delayed |
| **Muscle tone (hypotonia, ...)** | Mild movement coordination disorder | No | NA | Hypotonia |
| **Ataxia/spasticity** | No | No | NA | No |
| **Brain anomalies** | No (MRI normal) | NA | Diffuse cerebral atrophy on MRI; encephalopathy | Mild prominent cortical sulci denoting mild cortical atrophic changes and mild deep white-matter signal more than expected for age CT: no calcification |
| **Skeletal anomalies** | No | No | NA | No |
| **Recurrent infections** | No | No | NA | No |
| **Uric acid overproduction (gout, kidney stones, renal failure)** | Not known (no clinical signs) | NA | Minimal change nephrotic syndrome; GFR: 60ml/min/1,73m2 | Normal |
| **Serum uric acid** | Normal | NA | NA | Normal |
| **Urine (Hypoxanthine, Xanthine, uric acid levels)** | Quantitative amino acids in urine: Increased glycine excretion (may be nonspecific or indicate a mild renal tubular dysfunction) | NA | NA | Normal |
| **Other** | Nothing | No | Bilateral venous thrombosis, history of left-sided hemiparesis; ischemic stroke; Albumin 1.5g/l | Nothing |

**Table S5 Genotype and phenotype of individuals with CSMD2 *de novo* missense variants**
