## Supplementary material for "Haploinsufficiency, *de novo* and biallelic missense variants in *CSMD2* are associated with neurodevelopmental disorders": Table S6

| Number | 3.2 | 3.4 |
|---|---|---|
| Gender | M | F |
| Variant | CSMD1:NM_033225:c.5140C>T:p.R1714X | CSMD1:NM_033225:c.5140C>T:p.R1714X |
| Genotype | Homozygous | Homozygous |
| Variant inheritance | Inherited from mother (father deceased and untested) | Inherited from mother (father deceased and untested) |
| Date/Year of birth/Age | 26-30 yo | 31-35 yo |
| Onset | Early childhood | Early childhood |
| Intellectual disability | Yes | Yes |
| Learning and behavior | Delayed cognitive development | Delayed cognitive development |
| Motor development | Global development delay | Global development delay |

**Table S6 Genotype and phenotype of *CSMD1* homozygous stop-gain siblings**
